# A lyophilized open-source RT-LAMP assay for molecular diagnostics in resource-limited settings

**DOI:** 10.1101/2024.11.19.24317525

**Authors:** Martin Matl, Max J. Kellner, Felix Ansah, Irina Grishkovskaya, Dominik Handler, Robert Heinen, Benedikt Bauer, Luis Menéndez-Arias, Thomas O. Auer, Lucia L. Prieto-Godino, Josef M. Penninger, Vienna Covid-19 Detection Initiative (VCDI), Gordon A. Awandare, Julius Brennecke, Andrea Pauli

**Affiliations:** Research Institute of Molecular Pathology (IMP), Vienna BioCenter (VBC), Campus-Vienna-Biocenter 1, 1030 Vienna, Austria; Institute of Molecular Biotechnology of the Austrian Academy of Sciences (IMBA), Vienna BioCenter (VBC), Dr. Bohr-Gasse 3, 1030 Vienna, Austria; Vienna BioCenter PhD Program, Doctoral School of the University at Vienna and Medical University of Vienna, Vienna, Austria; Department of Laboratory Medicine, Medical University of Vienna, Austria; West African Center for Cell Biology of Infectious Pathogens (WACCBIP), College of Basic and Applied Sciences, University of Ghana, Legon, Ghana; Centro de Biología Molecular ‘Severo Ochoa’ (Consejo Superior de Investigaciones Científicas and Universidad Autónoma de Madrid), c/ Nicolás Cabrera 1, Campus de Cantoblanco-UAM, 28049 Madrid, Spain; Department of Biology, University of Fribourg, Fribourg, Switzerland; TReND in Africa, Brighton, UK; The Francis Crick Research Institute, London, UK; Helmholtz Centre for Infection Research, Braunschweig, Germany

## Abstract

A critical bottleneck for equitable access to population-scale molecular diagnostics is the limited availability of rapid, inexpensive point-of-care tests especially in low- and middle-income countries. Here, we developed an open-source *Reverse transcription loop-mediated isothermal amplification* (RT-LAMP) molecular assay for the detection of respiratory RNA virus infections. It is based on non-proprietary enzymes, namely HIV-1 reverse transcriptase, *Bst* LF DNA polymerase and UDG BMTU thermolabile uracil DNA glycosylase. Formulated as liquid or lyophilized reaction mixtures, these reagents enable sensitive colorimetric detection of respiratory samples without the need for prior nucleic acid isolation. We evaluated our open-source lyophilized RT-LAMP assay on clinical samples with suspected COVID-19 infection, demonstrating high sensitivity and 100% specificity compared to the gold standard *Reverse transcription quantitative polymerase chain reaction (*RT-qPCR). Reaction performance was unaffected by prolonged storage of lyophilized reagents at ambient or elevated temperatures. As a proof of concept, we evaluated the robustness and ease-of-use of lyophilized RT-LAMP reaction mixes through independent laboratory testing of COVID-19 samples in Ghana. Overall, our open-source RT-LAMP assay provides a flexible and scalable point-of-care test that can be adapted for rapid detection of various pathogens in resource-limited settings.

## INTRODUCTION

*In vitro* nucleic acid testing is an essential tool for containing outbreaks of infectious viral diseases (Peeling et al., 2022; Perkins et al., 2017). The COVID-19 pandemic has highlighted the unpreparedness of countries to rapidly establish testing programs, in part due to inadequate laboratory infrastructure to perform molecular testing (Mfuh et al., 2023). In addition, molecular diagnostic testing is commonly linked to the use of proprietary reagents, creating a dependency on commercial solutions. Although optimized to ensure robust performance, population-scale testing with commercial kits can be prohibitively expensive, especially for low- and middle-income countries that suffer from underinvestment in research and development (Petti et al., 2006). These countries also often have limited access to international supply chains for reagent distribution (Nkengasong, 2020; WHO, 2023). To address these challenges, several programs have been established to ensure equitable access to health products for all countries, such as the World Health Organization’s (WHO) COVID-19 Technology Access Pool (Venkatesan, 2023). However, key challenges remain to ensure the development and self-sufficient implementation of molecular testing programs for resource-limited settings (Jani & Peter, 2022).

Reverse transcription loop-mediated isothermal amplification (RT-LAMP) is a powerful molecular diagnostic tool with the potential to overcome many of the challenges of *in vitro* nucleic acid diagnostics (Feddema et al., 2024; Notomi et al., 2000). As an isothermal DNA amplification technique, RT-LAMP enables rapid and sensitive detection of RNA and DNA templates without the need for sophisticated laboratory infrastructure (Kellner et al., 2022; Notomi et al., 2000). Unlike conventional polymerase chain reaction (PCR), RT-LAMP assays are also compatible with colorimetric readouts (Goto et al., 2009; Scott et al., 2020; Tanner et al., 2015). Detection by visual color change can be particularly advantageous in resource-limited settings, as the binary reaction outcome (i.e., pathogen positive or negative) can be interpreted even by laypersons without specialized equipment.

Most commercially available RT-LAMP products to date contain proprietary, engineered enzymes and reaction components optimized to achieve high sensitivity and specificity (Barnes et al., 2021; Barnes Wayne et al., 2021; McMahon, 2021; Ong et al., 2012; Sharma et al., 2024). However, researchers and medical professionals in resource-limited settings often cannot afford these commercial solutions (2.9- 6$/reaction) or are located in countries without local distribution partners (Nkengasong, 2020; Ondoa et al., 2020). To overcome this, we and others have provided open-source protocols and reagents for low-cost RT-LAMP assays (Alekseenko et al., 2021; Bhadra et al., 2018; Kellner et al., 2022; Tomita et al., 2008). Nevertheless, easily deployable complete formulations, independent of cold chain storage of enzymes, reagents and oligonucleotide primers are urgently needed for point-of-care diagnostics and for monitoring applications in resource-limited settings (Jani & Peter, 2022). Here, we present a sensitive and robust, fully open access RT-LAMP reaction mix that meets all these criteria and allows colorimetric RT-LAMP from lyophilized reagents. Detailed protocols for preparing RT-LAMP enzymes and running assays are also provided on our website rtlamp.org.

## RESULTS

### Establishment of an effective autonomously produced open-source RT-LAMP reaction mixture

The performance of RT-LAMP assays is strongly dependent on the enzyme mix used, especially the type of reverse transcriptase (Kellner et al., 2022). To establish a low-cost, open-source RT-LAMP protocol for RNA detection, we therefore sought to determine which non-proprietary enzymes provide optimal reaction performance. We first compared several variants of the commonly used thermophilic DNA Polymerase from *Geobacillus stearothermophilus,* commercially available *Bst* LF (New England Biolabs), engineered *Bst* 2.0 DNA polymerase (NEB) and the improved DNA-binding and salt tolerant *Bst* 3.0 (NEB) against our in-house purified wild-type *Bst* LF (**Supplementary Figure 1**), Using a synthetic SARS-CoV-2 genomic RNA dilution series as input, we observed equal reaction specificity and sensitivity among all tested *Bst* polymerase variants at sample concentrations as low as 25-50 RNA copies/µl (**Figure 1A**). Of note, although the assay outcome after 35 minutes of RT-LAMP was identical, we observed a slightly faster onset of amplification in real-time monitored reactions for the engineered variants *Bst* 2.0 and *Bst* 3.0 compared to commercial *Bst* LF (**Supplementary Figure 2A**). Strikingly, the in-house produced *Bst* LF showed the best overall performance on synthetic SARS-CoV-2 genome standard using the ORF1ab-HMS LAMP primer set, which amplifies a region of the SARS-CoV-2 *ORF1ab* genic region (Rabe & Cepko, 2020) (**Figure 1A**, **Supplementary Figure 2A**).

**Figure 1:**
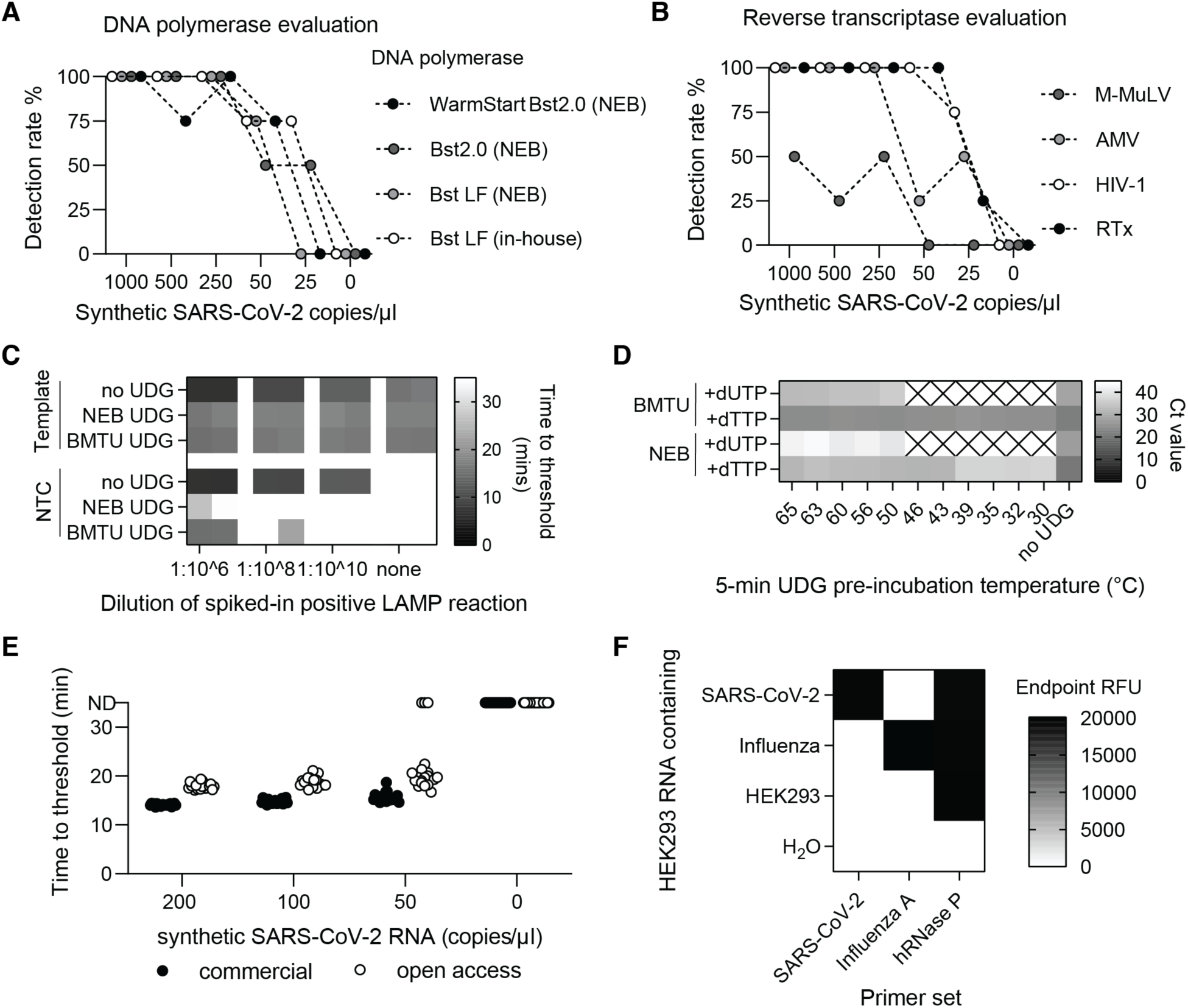
Assessing the performance of open-source RT-LAMP enzymes. (A) Comparison of DNA polymerase enzymes in RT-LAMP reactions. Detection rates of RT-LAMP reactions containing different DNA polymerases in addition to Warmstart RTx Reverse Transcriptase (NEB) and various concentrations of synthetic SARS-CoV-2 RNA. Results were obtained from four replicates per condition. (B) Comparison of Reverse Transcriptase enzymes in RT-LAMP reactions. Analogous to A) but for different Reverse Transcriptase enzymes in combination with in-house *Bst* LF DNA polymerase. M-MuLV = Moloney murine leukemia virus reverse transcriptase, AMV = avian myeloblastosis virus reverse transcriptase, and HIV-1 = human immunodeficiency virus 1 reverse transcriptase. (C) Cross-contamination prevention in RT-LAMP via uracil-DNA-glycosylase (UDG) enzymes. Diluted amounts of contaminating amplicons were added to RT-LAMP reactions containing different uracil-DNA-glycosylase (UDG) enzymes and synthetic SARS-CoV-2 template. No UDG enzyme and non-template control reactions were included. Shown are time-to-threshold values from real-time fluorescence RT-LAMP reactions performed in duplicates. In-house BMTU UDG enzyme was tested against Antarctic Thermolabile UDG (NEB). (D) Thermostability test of BMTU UDG and commercial UDG enzymes. In-house BMTU UDG and commercial Antarctic Thermolabile UDG (NEB) were pre-incubated at different temperatures for 5 minutes. UDG enzyme was then added to qPCR reactions targeting either dTTP-containing DNA template or dUTP containing DNA template. Cycles threshold values (Ct) from duplicate reactions are shown for each condition. Crossed box = not determined. (E) Limit of detection of open-access and commercial RT-LAMP reactions. Reactions prepared from in-house *Bst* LF, HIV-1 RT and BMTU UDG enzymes were compared to commercial reactions using the 2X WarmStart LAMP Kit (NEB) containing engineered proprietary enzymes. Synthetic SARS-CoV-2 RNA at defined copy numbers was used as template and 20 replicates were performed per condition. Time-to-threshold values from real-time fluorescence RT-LAMP reactions are shown. (F) Specificity test of RT-LAMP reactions detecting different pathogens. RT-LAMP reactions were assembled with *Bst* LF and HIV-1 RT. Primers targeting SARS-CoV-2, Influenza A or hRNaseP were tested against SARS-CoV-2 RNA, Influenza A RNA (each individually spiked into HEK 293 extracted RNA), HEK 293 extracted RNA alone and nuclease-free water as no-template control. Reactions were performed in 4 replicates and end-point relative fluorescent units are displayed.

Next, we determined which reverse transcriptase (RT) provides optimal reaction performance in combination with our in-house produced *Bst* LF polymerase. To this end, we compared open-source, in-house produced HIV-1 RT (**Supplementary Figure 1)**, Moloney Murine Leukemia Virus RT (MMuLV RT) and Avian Myeloblastosis Virus RT (AMV RT) with the commercially available engineered enzyme WarmStart® RTx (NEB) (**Figure 1B**). Of all the engineered RT enzymes, WarmStart® RTx provided the shortest onset of amplification (**Supplementary Figure 2B**) and showed the highest overall sensitivity (**Figure 1B)**. Strikingly, our in-house purified HIV-1 RT showed equal speed and sensitivity as WarmStart® RTx down to 25 RNA copies/µl (**Figure 1B, Supplementary Figure 2B**). We conclude that the thermostable HIV-1 RT is the enzyme of choice for open-source RT-LAMP assays.

Nucleic acid contamination is a serious risk associated with LAMP assays that can compromise test results and shut down entire diagnostic operations (Robinson-McCarthy et al., 2021). To reduce the risk of cross-contamination, we produced the thermolabile uracil DNA glycosylase from the psychrotrophic marine bacterium BMTU 3346 (hereafter referred to as BMTU UDG) (**Supplementary Figure 1)**, which is known to serve as an effective contamination prevention system in conjunction with dUTP for RT-LAMP assays (Hsieh et al., 2014; Jaeger et al., 2000; Kellner et al., 2022). Reaction performance in comparison to commercial Antarctic Thermolabile UDG (NEB) was tested in forced contamination experiments, where diluted amounts of dUTP containing SARS-CoV-2 LAMP amplicons were added to RT-LAMP reactions containing synthetic SARS-CoV-2 RNA. As expected, the absence of UDG resulted in indistinguishable amplification between positive and negative samples regardless of the amount of contaminating amplicon added (**Figure 1C**). In contrast, in-house produced or commercial thermolabile UDG added directly to the RT-LAMP reactions effectively prevented amplification in contaminated reactions in an amplicon concentration-dependent manner without substantially reducing reaction sensitivity (**Figure 1C, Supplementary Figure 2C**). Importantly, we confirmed that the prevention of contamination was based on the incorporation of dUTP, as dTTP-containing amplicons were amplified indiscriminately from the true RNA template regardless of the presence or absence of UDG (**Figure 1D**). Moreover, preincubation of in-house UDG above 50°C completely abolished its anti-contamination function, confirming the thermolabile nature of BMTU UDG (**Figure 1D**).

### Open-source RT-LAMP enables sensitive and specific RNA detection

To benchmark our complete open-source RT-LAMP reaction mix containing only in-house produced enzymes (HIV-1 RT, *Bst* LF and BMTU UDG) with the gold standard commercial RT-LAMP reagents from New England Biolabs, we used a dilution series of synthetic SARS-CoV-2 RNA. To establish a robust detection limit, RT-LAMP assays were performed in 20 replicates. The open-source RT-LAMP reagents resulted in a 100% (20/20) detection rate at 50 RNA copies/µl and an 85% (17/20) detection rate at 25 RNA copies/µl, which is comparable to the sensitivity measured for the commercial RT-LAMP reagents (**Figure 1E)**.

The sensitivity of the RT-LAMP assay can be further enhanced by a simple, 15-minute magnetic bead enrichment step that effectively concentrates the input nucleic acid material (**Supplementary Figure 2D**) (Kellner et al., 2022). We adapted this ‘bead-LAMP’ protocol to make it compatible with our open-source enzymes by (i) increasing the amount of polymerase present in the reaction mix, and (ii) using in-house manufactured nucleic acid capture beads (Oberacker et al., 2019). Bead-LAMP with open-source reagents on dilutions of synthetic SARS-CoV-2 RNA (Twist Bioscience) allowed reliable detection of 12.5 copies/µl (4/4 detected) and 3.125 copies/µl (3/4 detected) of the original sample concentration (**Supplementary Figure 2E**), demonstrating that the sensitivity of open-source RT-LAMP can be further increased with bead-LAMP. The specificity of our open-source RT-LAMP reaction mix was evaluated by adding either SARS-CoV-2 RNA, Influenza A RNA or water to purified RNA extracted from HEK293 cells. RT-LAMP assays were tested for SARS-CoV-2 (Rabe & Cepko, 2020), Influenza A (Takayama et al., 2019) or RNaseP targeting the human RNaseP transcript as an internal positive control (Broughton et al., 2020). Amplification was specific for each added RNA, and no cross-reactivity for either viral pathogen was observed (**Figure 1F)**.

Taken together, we have established an open-source RT-LAMP reagent mix that enables rapid, sensitive and specific colorimetric RNA detection and that performs on par with commercially available RT-LAMP reagents on contrived RNA samples. To enable low-cost and open distribution of the open-source enzymes, we have deposited *E. coli* expression vectors for His-tagged versions of these three wild-type enzymes with Addgene (www.addgene.org) and provide standard protocols for medium-scale protein expression and purification with this manuscript (Materials and Methods) and on our website (rtlamp.org).

### A sample inactivation procedure compatible with nucleic acid detection

In most approved nucleic acid diagnostic tests, RNA extraction is a necessary preanalytical step. However, RNA extraction is a bottleneck in the supply chain, adds a significant cost per sample, and is time- and labor intensive. Protocols that bypass the RNA extraction step have been reported previously (Fomsgaard & Rosenstierne, 2020; Myhrvold et al., 2018; Rabe & Cepko, 2020; Sandri et al., 2021; Ulloa et al., 2020). A 5-minute heat inactivation step using a commercially available QuickExtract™ DNA Extraction Solution (Lucigen) has been shown to result in sensitive SARS-CoV-2 nucleic acid detection, including for RT-qPCR and RT-LAMP (Joung et al., 2020; Kellner et al., 2022). We therefore aimed to develop an optimized, open-source, rapid sample inactivation protocol using readily available reagents to replace the proprietary QuickExtract™ solution.

The addition of the reducing agent tris(2-carboxyethyl)phosphine (TCEP) has been reported to be beneficial for preserving sample integrity after heat inactivation, especially of released naked RNA (Myhrvold et al., 2018; Rabe & Cepko, 2020). Therefore, we compared various TCEP-containing inactivation solutions with the commercial QuickExtract™ solution to develop a scalable, cost-effective alternative. Patient-derived SARS-CoV-2-positive nasopharyngeal swab and gargle samples were mixed with inactivation solutions and heat-inactivated at 95°C for 5 minutes (**Figure 2A)**. The inactivated samples were then stored at different temperatures (room temperature, 4°C and -20°C) and durations (0h, 4h, 16h) to simulate potential sample storage conditions. The integrity of viral RNA within the samples was assessed by direct-input RT-qPCR (Joung et al., 2020; Kellner et al., 2021). While heat-inactivated swab samples collected in Viral Transport Medium (VTM) without the addition of any inactivation buffer showed temperature-dependent degradation of viral RNA (**Figure 2B**), gargle samples in saline or HBSS proved more stable (**Figure 2C**), suggesting an intrinsically higher RNA stability in media lacking Fetal Bovine Serum (FBS). Although effective in preventing RNA degradation, the addition of TCEP resulted in sample heterogeneity and turbidity after the 5-minute heat-inactivation step, presumably from precipitated protein with a likely negative impact on RT-LAMP colorimetric detection (**Figure 2D**). Supplementation of the TCEP inactivation reagent with betaine and Proteinase K overcame this limitation, allowing for sensitive direct-input RT-qPCR and HNB colorimetric RT-LAMP without affecting performance (**Figure 2D, E, F**). Overall, this demonstrates that a TCEP/Betaine/Proteinase K inactivation solution enables rapid sample inactivation under conditions that preserve RNA integrity and maintain compatibility with downstream direct-input RT-qPCR and RT-LAMP.

**Figure 2:**
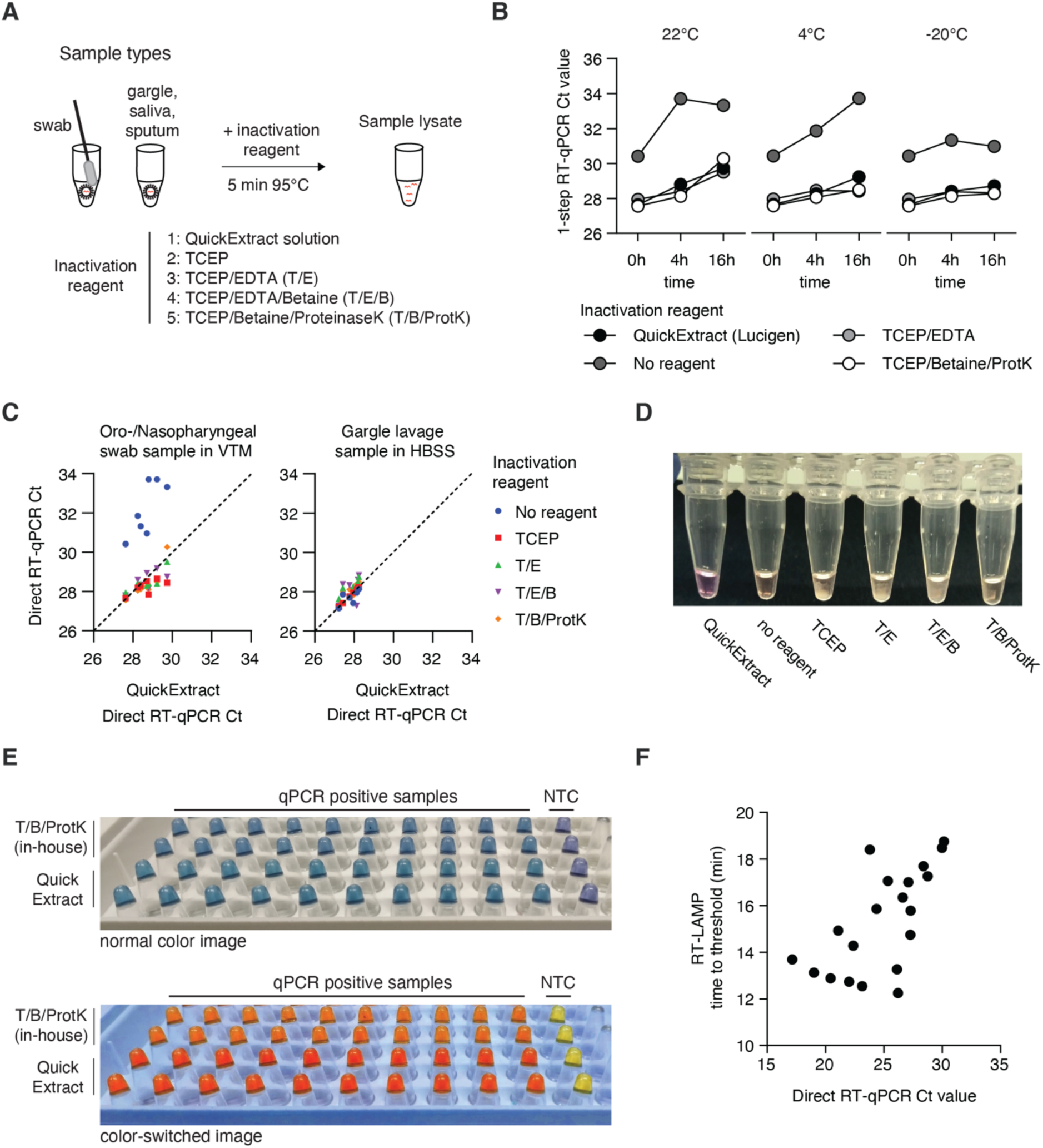
An optimized, quick inactivation solution for direct-input RT-LAMP and RT-qPCR. (A) Scheme illustrating quick sample inactivation. Patient-derived (self- or professionally collected) respiratory sample is inactivated by addition of an inactivation solution and heating for 5 minutes at 95°C. (B) Sample stability over time with different inactivation reagents. A mock sample in viral transfer medium was inactivated by adding inactivation reagents and heating for 5 minutes at 95°C. Samples were tested with one-step direct-input RT-qPCR at 0, 4 and 16 hours of storage at room temperature (left), 4°C (middle) and -20°C (right). (C) Comparison of two commonly used respiratory tract samples. Sample stability after inactivation with different solutions was determined for independent swab samples in VTM or gargle lavage in HBSS. Ct-values of open-source inactivation solutions were compared to QuickExtract^TM^ solution. (D) Image demonstrating turbidity generated upon heat-inactivation of VTM samples with different inactivation solutions. Smartphone image of mock nasopharyngeal sample in VTM taken after the addition of inactivation solutions and 5-minute incubation at 95°C. (E) Colorimetry images of RT-LAMP reactions prepared after inactivation of samples with QuickExtract^TM^ or in-house inactivation solution. Inactivated nasopharyngeal swab and gargle samples were tested by open access RT-LAMP to assess the HNB-colorimetric readout. (F) Comparison of time to threshold values for RT-LAMP and cycle to threshold for RT-qPCR using direct input samples. Time to threshold in minutes is reported for RT-LAMP whereas cycle threshold is reported for RT-qPCR. As evident from the poor correlation, quantitative measurement of target copy number is not possible via RT-LAMP.

#### Lyophilization of open-source RT-LAMP enzymes and reaction mixes

Having established a direct-input RT-LAMP assay using only open-source reagents, we next sought to develop a lyophilized mixture to enable global distribution independent of cold chain logistics. We established two separate lyophilization protocols using glycerol-free open-source RT-LAMP enzymes and D-trehalose as a cryoprotectant (Carter et al., 2017): A lyo-RT-LAMP enzyme mix containing only open-source enzymes (‘enzyme mix’), which upon reconstitution provides a 20x concentrated glycerol enzyme mixture for immediate use or storage at -20°C. We also developed a lyo-RT-LAMP reagent mix containing enzymes, dNTPs and primers for immediate use (‘reagent mix’) (**Figure 3A**) (see Materials and Methods and rtlamp.org for details).

**Figure 3:**
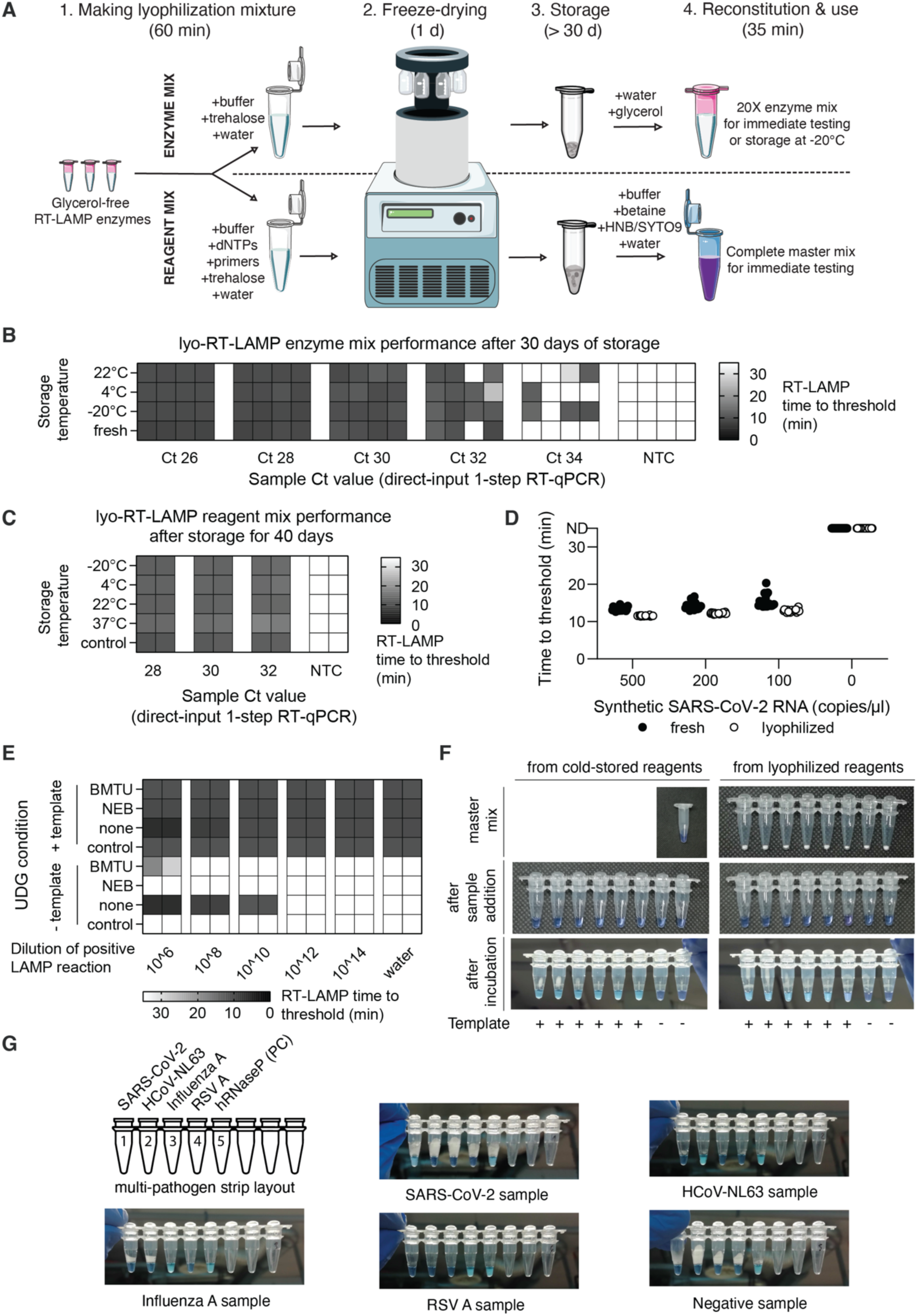
An open-source lyophilized RT-LAMP mixture. (A) Schematic representation of lyophilization options for freeze-drying RT-LAMP enzymes and reagents. Glycerol free enzymes can be lyophilized either as a concentrated enzyme mix or as a reagent mixture with dNTPs and primers. Trehalose is used in both options as a cryoprotectant. Mixtures are flash-frozen in liquid nitrogen, lyophilized using a freeze-dryer and stored in dry conditions at various temperatures for transport. Before use, lyophilized mixtures are reconstituted. (B) Performance of reactions assembled with reconstituted RT-LAMP enzyme mix. RT-LAMP enzyme mix was lyophilized and stored at 22°C, 4°C and -20°C. After 30 days, lyophilized enzymes were reconstituted and used to assemble RT-LAMP reactions; control RT-LAMP reactions were prepared with non-lyophilized enzymes stored at -20°C. Reactions were tested on a four-fold, five-step dilution series of a positive sample in four replicates by RT-LAMP, recording the time-to-threshold for each reaction. In addition, the sample dilutions were measured by direct-input RT-qPCR, and Ct values are displayed under the columns. (C) Time to threshold for reactions prepared using a reconstituted RT-LAMP reagent mix. RT-LAMP reagent mix was lyophilized and stored at -20°C, 4°C, 22°C and 37°C. After 40 days, RT-LAMP reagents were reconstituted using the reconstitution buffer and reactions were tested on a four-fold, three-step dilution series of a positive sample in duplicates by RT-LAMP, recording the time-to-threshold for each reaction. In addition, the sample dilutions were measured by direct-input RT-qPCR, and Ct values are displayed under the columns. (D) Sensitivity comparison of RT-LAMP reactions assembled from lyophilized and non-lyophilized RT- LAMP reagents. Lyophilized and freshly prepared reactions were compared in 20 replicates using a dilution of SARS-CoV-2 synthetic RNA (Twist Biosciences) to assess the detection limit. Time to threshold values are shown. (E) Forced contamination experiment using lyophilized RT-LAMP reagents. In-house BMTU UDG enzyme, commercially available Antarctic Thermolabile UDG (New England Biolabs) and no UDG were included in three respective RT-LAMP reagent mixes for lyophilization. After freeze-drying, the reagent mixes were reconstituted into RT-LAMP master mixes and compared to freshly prepared reactions in a forced contamination experiment as described earlier (Figure 1 – C). (F) HNB colorimetric readout is compatible with lyophilized RT-LAMP reagent mixture. Reactions were prepared in parallel from cold-stored enzymes (left) and lyophilized reaction mix (right) to compare colorimetric readout. Smartphone images were taken after lyophilization/preparation of master mix (top row), after addition of sample (middle row) and after a 35-minute incubation at 63°C (bottom row) to showcase the color change. (G) Proof-of-concept multi-pathogen respiratory virus RT-LAMP test. An 8-well PCR strip was filled with singe-reaction aliquots of RT-LAMP reagent mix for different target RNAs and freeze-dried. A multi-pathogen test strip targeting SARS-CoV-2, human coronavirus NL63 (HCoV-NL63), Influenza A, Respiratory Syncytial Virus A (RSV A) and human RNaseP (PC, positive control) was prepared from lyophilized reagents. Shown are the HNB RT-LAMP colorimetric results after reconstitution and addition of mock respiratory sample containing the respective pathogen RNA. A light blue color indicates a positive result.

To measure the performance of lyophilized enzymes after long-term storage, we reconstituted the freeze-dried lyo-RT-LAMP mixtures stored for 10 and 30 days at different temperatures (-22°C, 4°C, room temperature) and compared the reaction sensitivity to a freshly prepared enzyme mixture using a contrived, rapidly inactivated dilution series of a SARS-CoV-2 positive sample. We observed little to no measurable loss of sensitivity and no difference in reaction onset (time-to-threshold) in RT-LAMP reactions prepared with reconstituted enzymes compared to fresh reagents even after 30 days of storage (**Figure 3B; Supplementary Figure 3A**). Similar results were obtained for the lyophilized ‘reagent-mix’, with no change in reaction performance after 10 and 40 days of storage at different temperatures for contrived crude SARS-CoV-2 or synthetic SARS-CoV-2 RNA dilutions (**Figure 3C, D; Supplementary Figure 3B)**. In addition, a cross-contamination test showed that BMTU UDG did not lose activity during lyophilization and was able to protect against cross-contamination in this temperature-stable formulation (**Figure 3E**).

To demonstrate the feasibility of providing open-source reagent mixes for home and remote point-of-care settings, we prepared individually lyophilized reagent mixes in PCR strips (1 reaction/tube) (**Figure 3F**). Such a lyophilized PCR strip with reagent mix was reconstituted with reconstitution buffer and tested in parallel with an RT-LAMP mix prepared from cold stored enzymes (stored in glycerol-containing storage buffer at -20°C). The strips were tested on the same sample, demonstrating robust colorimetric reaction performance in lyophilized and cold-stored RT-LAMP reagent mixes **(Figure 3F).**

Finally, we aimed to demonstrate the utility of lyo-RT-LAMP reagent mixes for the detection of viral pathogens beyond SARS-CoV-2. In a proof-of-concept experiment, we assembled a multi-pathogen respiratory virus RT-LAMP test for rapid testing at the point-of-care, targeting SARS-CoV-2, human coronavirus NL63 (Amsterdam I), Influenza A virus (H1N1 A/WSN/1933), Respiratory Syncytial Virus (RSV A2), and human RNaseP in a single 8-well PCR strip containing lyo-RT-LAMP reagent mixes with primer sets specific to these viruses. The multi-pathogen lyophilized RT-LAMP strips were reconstituted by the addition of reconstitution buffer and tested on contrived samples containing individual virus RNA samples in a quick-inactivated negative control sample. The resulting visible color change revealed a clear and pathogen-specific nucleic acid amplification (**Figure 3G**), demonstrating the broad applicability of this simple solution for point-of-care testing. Overall, we have established two versatile lyophilization solutions for open-source RT-LAMP reagents that circumvent the current cold chain dependency of SARS-CoV-2 molecular test kit distribution and showed their potential to detect other viruses.

#### Performance of direct-input open-source RT-LAMP on clinical samples

To determine the sensitivity and specificity of lyophilized open-source RT-LAMP reagents on clinical samples, two validation studies were conducted, one in Vienna, Austria and another one in Accra, Ghana. In Vienna, a total of 192 patient respiratory tract specimens (103 oro-/nasopharyngeal swabs in VTM and 89 gargle samples in Hank’s Balanced Salt Solution (HBSS) or saline solution (0.9% NaCl)) were heat-inactivated in home-made complete inactivation solution and tested with RT-LAMP using lyophilized open access RT-LAMP reagent mix presented in this study. In parallel, samples were measured by direct-input RT-qPCR for reference and to obtain quantitative Ct values. 80 out of the 192 samples were identified as SARS-CoV-2 positive by direct-input RT-qPCR for the SARS-CoV-2 N-gene using the CDC N1 primer set. Direct input lyophilized open-source RT-LAMP reactions showed a weak but detectable negative correlation of the concentration of viral RNA in patient samples with the time-to-threshold value for real-time fluorescence RT-LAMP reactions and resulted in robust HNB-based color changes for positive patient samples, indicating complete amplification after a run time of 35 minutes (**Figure 4A, B**). The lyo-RT-LAMP assay correctly detected 52 out of the 80 samples identified as positive by RT-qPCR (overall sensitivity of 65.0% (95% CI: 54.1 – 75.6%)), with a 50% limit of detection (LOD50) for a sample with a Ct-value of 32.53 (95% C.I: 31.34-33), corresponding to approximately 100 copies per reaction (**Figure 4C, Supplementary Figure 4A**). All 112 RT-qPCR negative samples were also identified as negative by lyo-RT-LAMP, giving a specificity of 100% (**Figure 4D, Supplementary Figure 4B)**.

**Figure 4:**
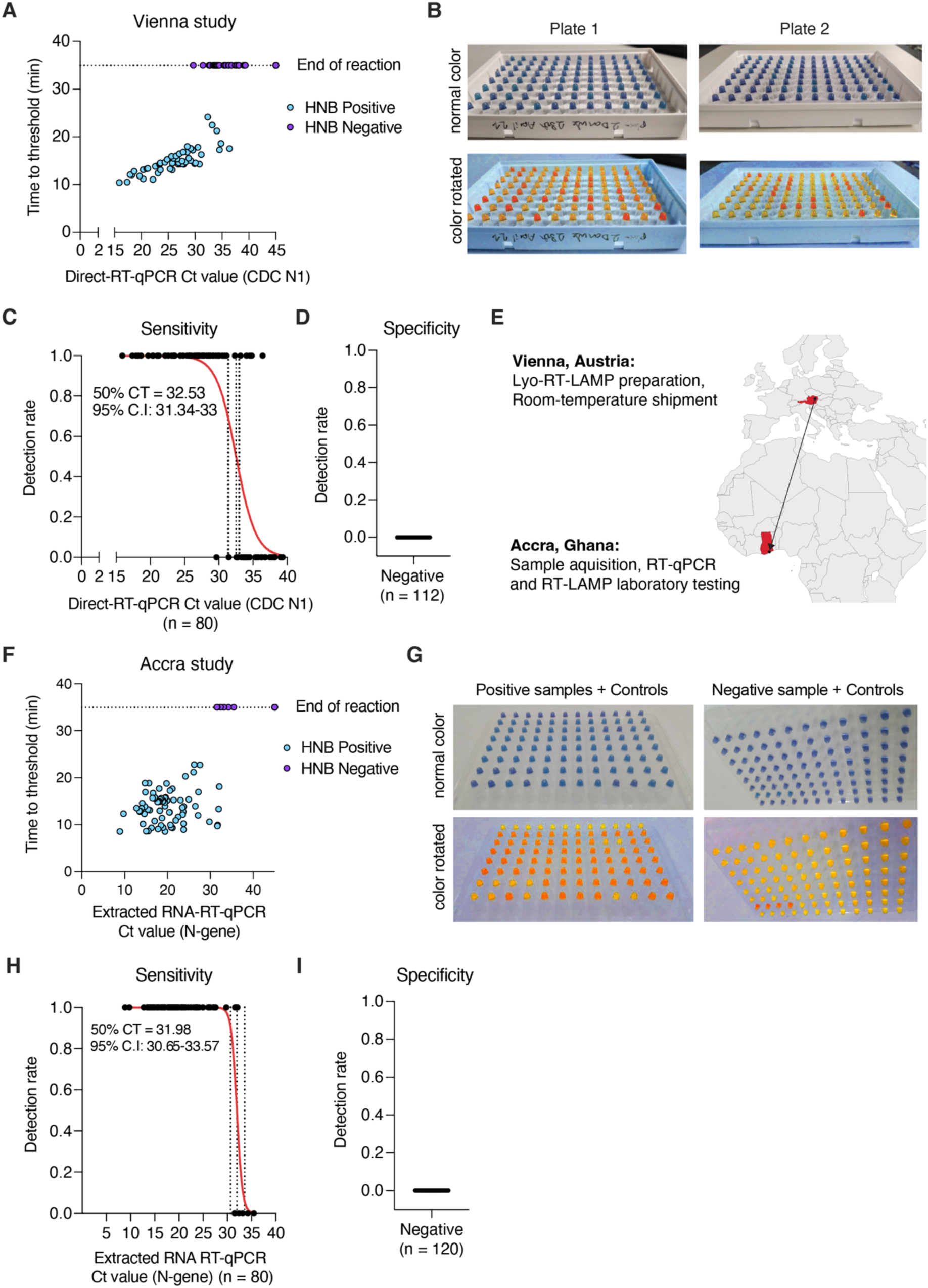
Performance evaluation of lyophilized RT-LAMP reagent mix on clinical specimens in Austria and Ghana. (A) Performance of direct-input open-source RT-LAMP on clinical samples in Vienna. 192 samples with suspected COVID-19 infection were inactivated and tested in parallel with RT-qPCR targeting the N-gene, or HNB colorimetric RT-LAMP from lyophilized reagents. Shown are the time-to-threshold values obtained from real-time RT-LAMP reactions in comparison to RT-qPCR Ct-values. Each dot represents an individual sample. The color of each dot indicates the colorimetric results from HNB RT-LAMP reactions. (B) Colorimetric HNB RT-LAMP results from samples shown in A). Raw (top) or color-converted (bottom) images are shown. (C) Sensitivity of lyophilized RT-LAMP reactions. Each dot represents a single sample. A simple logistic regression analysis was performed for samples containing different amounts of viral RNA (measured via RT-qPCR) and plotted as red line. 95% confidence intervals and 50% detection limits are indicated as dashed lines and added as textbox. (D) Specificity of lyophilized RT-LAMP reactions. Each dot represents a single sample. RT-LAMP detection rates for true-negative samples determined by RT-qPCR are shown. (E) Clinical sample validation performed in Vienna, Austria, and Accra, Ghana. Lyophilized reagents were prepared in Vienna and shipped to Accra. Each site performed an independent validation study. (F) Performance of direct-input open-source RT-LAMP on clinical samples in Accra, Ghana. RNA from 192 samples with suspected COVID-19 infection was extracted and tested in parallel with RT-qPCR targeting the N-gene, or HNB colorimetric RT-LAMP from lyophilized reagents. Shown are the time-to- threshold values obtained from real-time RT-LAMP reactions in comparison to RT-qPCR Ct-values. Each dot represents an individual sample. The color of each dot indicates the colorimetric results from HNB RT-LAMP reactions. (G) Colorimetric HNB RT-LAMP results from samples shown in (F). Raw (top) or color-converted (bottom) images are shown. (H) Sensitivity of lyophilized RT-LAMP reactions. Each dot represents a single sample. A simple logistic regression analysis was performed for samples containing different amounts of viral RNA (measured via RT-qPCR) and plotted as red line. 95% confidence intervals and 50% detection limits are indicated as dashed lines and added as textbox. (I) Specificity of lyophilized RT-LAMP reactions. Each dot represents a single sample. RT-LAMP detection rates for true-negative samples determined by RT-qPCR are shown.

#### Performance on clinical specimens at an independent laboratory in Ghana

To demonstrate the cold chain independence, robust deployability and usability of our assay, we shipped lyophilized RT-LAMP reagent mixes to the West African Center for Cell Biology of Infectious Pathogens (WACCBIP) in Accra, Ghana (**Figure 4E**). A total of 200 respiratory tract (sputum) specimens with suspected SARS-CoV-2 infection were tested in this study. Samples were processed by a 5-minute heat inactivation with home-made complete inactivation solution; in parallel, RNA extraction followed by RT-qPCR targeting the N-gene was performed as a reference. RT-qPCR identified 80 out of 200 samples as positive for SARS-CoV-2. Of these, 73 and 72 samples were detected as positive by real-time fluorescence and colorimetric lyophilized RT-LAMP, respectively (**Figure 4F, G**). The sensitivity of the assay on this sample set was 91.3% and 90.0% for direct input samples, as measured by real-time fluorescence and colorimetry, respectively, with a LOD_50_ for samples with a Ct-value of 31.98 (95% C.I: 30.65-33.57) (**Figure 4H)**. The specificity for both sample input types and both readouts was 100%, as no false positives were detected in either fluorescence or colorimetry (**Figure 4I)**. These data demonstrate that our established open-source lyophilized RT-LAMP reagent mix can be deployed at room temperature to remote locations with no apparent loss of reaction performance.

## DISCUSSION

Here, we addressed two grand challenges in the development of equitable access to molecular testing: 1) Establishment of open-source assays for cost-effective nucleic acid detection at scale, and 2) a lyophilized assay formulation for cold chain-independent shipment and storage. We present a rapid, accurate and sensitive molecular RT-LAMP test for SARS-CoV-2 detection and beyond, using non-proprietary enzymatic components that can be produced and lyophilized in most laboratory settings. Due to its isothermal colorimetric nature, this nucleic acid amplification technique is especially suited for resource-limited settings. To ensure open access, we have made bacterial expression plasmids available through Addgene (plasmid IDs 159148 (*Bst* LF), 159149 (HIV-RT) and 172197 (BMTU UDG)) and established a website (rtlamp.org) with detailed protocols for autonomous production of reaction components and setup of RT-LAMP assays. Using these expression plasmids, a bacterial culture of only four liters yields enough recombinant protein to perform 200.000 individual tests. This study presents expression and purification protocols for all three open-source enzymes critical for sensitive and specific RT-LAMP, but further steps could be taken to make the purification protocols as low-tech as possible. Availability of FPLC systems and purification columns may be an issue for some laboratories, so optimizing these protocols or even redesigning expression constructs to work with simpler or even single-step purification methods could be of practical use (as reviewed by Leonhardt et al., 2023).

To eliminate the need for a dedicated RNA extraction step for nucleic acid diagnostics, we have tested and optimized a non-proprietary home-made inactivation solution for use with colorimetric RT-LAMP and RT-qPCR with a variety of respiratory sample types, including oro-/nasopharyngeal swabs, gargle lavage and sputum. The final inactivation reagent, formulated at 10x concentration and containing TCEP, Betaine and Proteinase K, allows for maximum sample volume input without compromising the test performance. In particular, the use of the inactivation solution prevents protein precipitation in the sample, which otherwise strongly affects the HNB-based colorimetric readout. The choice of HNB as a colorimetric indicator enables the use of sample types with varying pH values, which is not possible using phenol red dye. A possible drawback of HNB is its somewhat weaker color change, however, this can be mitigated using simple image enhancement tools, also available on *colorimetry.net* (Kellner et al., 2022). With our optimized RT-LAMP enzyme mixture containing *Bst* LF, HIV-RT and BMTU UDG, direct-input colorimetric RT-LAMP showed excellent sensitivity and 100% specificity using contrived and clinical respiratory samples.

Importantly, we were able to establish protocols for lyophilized enzyme-only (*Bst* LF, HIV-RT, BMTU UDG) and complete RT-LAMP reagent mixtures (enzymes, dNTPs, oligonucleotide primers). The fully open-source lyo-RT-LAMP reagents were stable after storage at ambient or elevated temperatures (37°C) for at least 40 days without negatively affecting reaction performance. Future experiments are needed to determine whether other laboratory sites will be successful in producing lyophilized reagents according to our protocol, and how other environmental or experimental factors, such as humidity and varying degrees of practical molecular biology skills influence the assay’s sensitivity and specificity.

Finally, we benchmarked the performance of open-source lyophilized RT-LAMP reagents using clinical samples with suspected COVID-19 infection. The two studies, conducted independently in Vienna, Austria, and Accra, Ghana, showed high concordance in terms of assay sensitivity and specificity. In fact, lyophilized reagents produced equivalent reaction results compared to cold chain stored components, suggesting that our lyophilized reagent mixture retains full functionality in RT-LAMP assays. Our limited experiments with other common respiratory viruses further demonstrate the applicability of open-source lyophilized RT-LAMP reagent mixtures for use *in vitro* nucleic acid detection beyond SARS-CoV-2.

Taken together, we have established a simple and cost-effective solution for nucleic acid detection based on non-proprietary enzymes for point-of-care diagnostics in resource-limited settings. Our protocols also enable researchers and institutions to autonomously produce reagents and perform testing in a decentralized setting. Ultimately this should help to provide equitable access to pathogen surveillance and epidemic management in various settings around the world.

## MATERIALS AND METHODS

### Expression and purification of open access RT-LAMP enzymes

Expression plasmids for *Bst* LF, HIV-RT and BMTU UDG were generated in pET expression vectors by Gibson cloning, using synthetic DNA coding for the enzymes as ordered as gBlocks (IDT). Expression plasmids are available through Addgene: plasmid IDs 159148 (*Bst* LF), 159149 (HIV-RT) and 172197 (BMTU UDG). *E. coli* BL21(DE3) bacterial cells were transformed with the respective pET-series expression plasmids and overnight cultures were grown at 37°C in LB medium at 180 rpm in the presence of ampicillin. Large scale expression was performed in the ZYP-5052 autoinduction medium (Studier, 2005). The cells were grown for 5 hours 30 minutes at 37°C, followed by overnight expression at 18°C. The cells were then harvested by centrifugation (4000 x g, 15 min) and stored at -80°C.

*Bst* LF purification was carried out as follows: The cells were resuspended in the lysis buffer (20 mM Tris-HCl pH 7.5, 500 mM NaCl, 0.5 mM TCEP and Benzonase^®^). Cell lysis was accomplished using a single cycle in a cell disruptor (Constant systems Ltd), with the pressure set to 1.4 kBar. Lysates were clarified by centrifugation (42.000 x g, 45 min, 4°C), and a two-step purification protocol using an ÄKTA Protein Purification System (GE Healthcare Life Sciences) at 8°C was then followed to obtain the recombinant protein. First, the supernatant was applied to the His-Trap Crude 5 ml column (GE Healthcare Life Sciences) previously equilibrated with the lysis buffer. The column was washed with 20 column volumes of the lysis buffer, containing 20 mM imidazole. Then, the protein was eluted using a step gradient of imidazole (equilibration buffer with 250 mM imidazole and then with 500 mM imidazole). Fractions were examined by SDS-PAGE for protein content and purity and pooled according to the presence of *Bst* LF and diluted with the RQA buffer containing 20 mM Tris-HCl pH 7.5, 0.5 mM TCEP to a final concentration of 50 mM NaCl. As a second step, the diluted fractions were applied to Resource Q 6 ml ion exchange column (GE Healthcare Life Sciences) pre-equilibrated with the RQA buffer. The protein was then eluted using a linear gradient of 0.05 to 0.74 M NaCl in the RQA buffer. The peak fractions were combined and dialyzed overnight against storage buffer consisting of 40 mM Tris-HCl pH 7.5, 50 mM NaCl, 1 mM DTT and 10% glycerol, flash frozen in liquid nitrogen and stored at -80°C until use.

HIV-1 RT purification was carries out as follows: The cells were resuspended in the lysis buffer (20 mM Tris-HCl pH 7.5, 500 mM NaCl, 0.5 mM TCEP and Benzonase^®^). Cell lysis was accomplished using a single cycle in a cell disruptor (Constant systems Ltd), with the pressure set to 1.4 kBar. Lysates were clarified by centrifugation (42.000 x g, 45 min, 4°C), and a three-step purification protocol using an ÄKTA Protein Purification System (GE Healthcare Life Sciences) at 8°C was then followed to obtain the recombinant protein. First, the supernatant was applied to the His-Trap Crude 5 ml column (GE Healthcare Life Sciences) previously equilibrated with the lysis buffer. The column was washed with 20 column volumes of the lysis buffer, containing 20 mM imidazole. Then, the protein was eluted using a step gradient of imidazole (equilibration buffer with 250 mM imidazole and then with 500 mM imidazole). Fractions were examined by SDS-PAGE for protein content and purity and pooled according to the presence of HIV-1 RT and diluted with the RSA buffer containing 20 mM Tris-HCl pH 7.5, 0.5 mM TCEP to a final concentration of 50 mM NaCl. As the second step, diluted fractions were applied to Resource S 6 ml ion exchange column (GE Healthcare Life Sciences) pre-equilibrated with the RSA buffer. The protein was then eluted using a linear gradient of 0.05 to 0.74 M NaCl in the RQA buffer. For the third step of the purification protocol, peak fractions were combined and applied to HiLoad 16/600 Superdex 200 pg column (GE Healthcare Life Sciences) equilibrated with the storage buffer consisting of 50 mM Tris-HCl pH 7.5, 50 mM NaCl, 0.5 mM TCEP and 10% glycerol. Eluted fractions were analyzed by SDS-PAGE, pooled according to the protein purity, concentrated, flash frozen in liquid nitrogen and stored at -80°C until use.

BMTU 3346 UDG purification was carried out as follows: The cells were resuspended in the lysis buffer (20 mM Tris-HCl pH 8.0, 500 mM NaCl, 0.5 mM TCEP and Benzonase^®^). Cell lysis was accomplished using a single cycle in a cell disruptor (Constant systems Ltd), with the pressure set to 1.4 kBar. Lysates were clarified by centrifugation (42.000 x g, 45 min, 4°C), and a two-step purification protocol using an ÄKTA Protein Purification System (GE Healthcare Life Sciences) at 8°C was then followed to obtain the recombinant protein. The supernatant was applied to the His-Trap Crude 5 ml column (GE Healthcare Life Sciences) previously equilibrated with the lysis buffer. The column was washed with 20 column volumes of the lysis buffer, containing 20 mM imidazole. Then, the protein was eluted using a step gradient of imidazole (equilibration buffer with 250 mM imidazole and then with 500 mM imidazole). Fractions were examined by SDS-PAGE for protein content and purity and pooled according to the presence of *Bst* LF and diluted with the RQA buffer containing 20 mM Tris-HCl pH 8.0, 0.5 mM TCEP to a final concentration of 50 mM NaCl. Diluted fractions were applied to Resource Q 6 ml ion exchange column (GE Healthcare Life Sciences) pre-equilibrated with the RQA buffer. The protein was then eluted using a linear gradient of 0.05 to 0.74 M NaCl in the RQA buffer. The peak fractions were combined and dialysed overnight against storage buffer consisting of 20 mM Tris-HCl pH 8.0, 50 mM NaCl, 0.5 mM TCEP flash frozen in liquid nitrogen and stored at -80°C until use.

#### Assembly of RT-LAMP reactions to identify open-source enzymes

Reactions comparing different RT-LAMP enzymes were carried out in 1X Isothermal Amplification Buffer (20 mM Tris-HCl pH 8.8, 10 mM (NH_4_)_2_SO_4_, 50 mM KCl, 2 mM MgSO_4_, 0.1% Tween® 20), 1.4 mM dNTP mix (25 mM stock, Larova), 0.7 mM dUTP (100 mM stock, Larova), 6 mM MgSO_4_ (8 mM final including magnesium sulfate from buffer, NEB), 0.4 M betaine (Sigma-Aldrich), 2 µM SYTO™ 9 fluorescent dye (100 µM stock in DMSO, ThermoFisher), 120 µM hydroxynaphthol blue trisodium salt (from 3 mM stock in nuclease-free water, Hach), oligonucleotide primers from the HMS-1 primer set (Rabe & Cepko, 2020) at final concentrations of 0.2 μM for F3/B3, 0.4 μM for LB/LF and 1.6 μM FIP/BIP (Sigma-Aldrich) and nuclease-free water to a total volume of 8 µl, reserving 2 µl for sample volume and volume for enzymes, see below.

To test different DNA polymerases, WarmStart® RTx Reverse Transcriptase (NEB) was added at 0.3 U/µl and Antarctic Thermolabile UDG (NEB) was added at 0.02 U/µl to all reactions. DNA polymerases were added to individual reactions at final concentrations according to manufacturer’s instructions, namely: 0.32 U/µl *Bst* DNA Polymerase Large Fragment (NEB), 0.32 U/µl *Bst* 2.0 DNA Polymerase (NEB), 0.32 U/µl *Bst* 2.0 WarmStart® DNA Polymerase (NEB), 0.32 U/µl *Bst* 3.0 DNA Polymerase (NEB) while *Bst* LF DNA polymerase (in-house produced) was used at the empirically determined optimal concentration of 20 ng/µl.

To test different reverse transcriptases, in-house *Bst* LF DNA polymerase was added at a final concentration of 20 ng/µl and Antarctic Thermolabile UDG (NEB) was added at 0.02 U/µl to all reactions. Reverse transcriptases were added according to manufacturer’s instructions at the final concentrations of 0.2 U/µl AMV Reverse Transcriptase (NEB), 4U/µl M-MuLV Reverse Transcriptase (NEB), 0.3 U/µl WarmStart® RTx Reverse Transcriptase (NEB) while HIV-1 reverse transcriptase (in-house produced) was used at the empirically determined concentration of 7.5 ng/µl.

Reactions with different DNA polymerases and reverse transcriptases were tested by the addition of synthetic SARS-CoV-2 RNA (Twist Biosciences) diluted in nuclease-free water to a concentration of 1000, 500, 250, 50, 25 and 0 copies/µl in the sample and incubation at 63°C for 60 minutes, with real-time fluorescence reading in SYBR/FAM wavelength every minute in a BioRad CFX96 instrument. Reactions were performed in 4 replicates.

To test different uracil-DNA-glycosylase enzymes, reactions were prepared as stated above, differing in the enzymes and primers used. LAMP ORF3a-A primer set from (Schermer et al., 2020) were used exclusively in this experiment in order to avoid causing contamination for the primer pair used in the rest of the study. In-house *Bst* LF DNA polymerase and in-house HIV-1 reverse transcriptase were added at a final concentration of 20 ng/µl and 7.5 ng/µl, respectively, to all reactions. A pre-contamination reaction was set up with the LAMP ORF3a-A primer set and a SARS-CoV-2 positive sample was amplified by incubation at 63°C for 30 minutes. The resulting positive reaction was serially diluted in a 1:100 manner, diluting the positive reaction to 1:10^2^, 1:10^4^, 1:10^6^, 1:10^8^, 1:10^10^, respectively. RT-LAMP master mixes were prepared with LAMP ORF3a-A primer set and HIV-1 RT and *Bst* LF DNA polymerase as described above. For the commercial UDG condition, Antarctic Thermolabile UDG (NEB) was added to the reactions in the final concentration of 0.02 U/µl, for the BMTU UDG condition, in-house purified BMTU UDG was added to the reactions in the final concentration of 0.025 ng/µl and for the no UDG condition UDG storage buffer was added to the reactions to match the volume of added UDG in the previous conditions. Positive inactivated SARS-CoV-2 sample was added to half of the reactions (+ template), and negative sample was added to the second half (- template), to distinguish between true positive and false positive in the presence of amplicons. Reactions were spiked with the contaminant dilutions and incubated at 63°C for 35 minutes. The reactions were performed in duplicates.

To test the thermostability of UDG enzymes using a qPCR-based assay, two dsDNA template amplicons were prepared (i) a 0.4 kb dsDNA amplicon generated via PCR with equimolar ratios of dTTP and dUTP in the reaction and (ii) 1 kb dsDNA amplicon generated via PCR with no dUTP present in the reaction. These are the dUTP and dTTP target, respectively. Two separate PCR master mixes were prepped to amplify the two respective targets, the reaction consisting of 1X Standard Taq Reaction Buffer (NEB), 250 µM dNTPs each (including dUTP), 0.1 µM forward primer, 0.1 µM reverse primer, 1 µM SYTO-9™ intercalating fluorescent dye, 0.25 U *Taq* DNA polymerase, and template dsDNA (0.01 ng/µl) to a final reaction volume of 20 µl, reserving. 0.4 µl for addition of UDG enzyme. In-house expressed and purified BMTU UDG enzyme (2.5 ng/µl) and Antarctic Thermolabile UDG (NEB) were incubated in a thermocycler on a gradient setting (30°C – 65°C) for 5 minutes. After incubation, UDG enzymes were immediately put on ice and 0.4 µl of enzyme was added to each PCR reaction on ice. PCR was performed in a BioRad CFX qPCR cycler with the following thermal cycling settings: denaturation 96°C for 30 s, annealing 60°C for 30 s, elongation 72°C for 60 s, repeated for a total of 45 cycles. Fluorescence readings in the FAM/SYBR range were taken after every elongation cycle.

#### Open-source RT-LAMP and bead-LAMP assays

RT-LAMP master mix consists of 1X Isothermal Amplification Buffer (20 mM Tris-HCl pH 8.8, 10 mM (NH_4_)_2_SO_4_, 50 mM KCl, 2 mM MgSO_4_, 0.1% Tween® 20, entire buffer prepared from stock solutions as a 10X concentrate or purchased from NEB, stored at -20°C), 1.4 mM dNTPs each (25 mM each, stock solution in ultrapure water, Larova), 0.7 mM dUTP (100 mM stock in ultrapure water, Larova), 6 mM MgSO_4_ (8 mM final including magnesium sulfate from Isothermal Amplification Buffer, from 1M stock solution or 100 mM stock solution NEB), 0.4 M betaine (from 5M stock solution prepared from powder, solution stored at -20°C, Sigma-Aldrich), 2 µM SYTO™ 9 fluorescent dye (100 µM stock solution in DMSO, for performing fluorescence readout, ThermoFisher), 120 µM hydroxynaphthol blue (HNB) trisodium salt (from 3 mM stock in nuclease-free water, stored at 4°C or -20°C, Hach), oligonucleotide primers from the HMS-1 primer set for SARS-CoV-2 detection (Rabe & Cepko, 2020) at final concentrations of 0.2 μM for F3/B3, 0.4 μM for LB/LF and 1.6 μM FIP/BIP (resuspended in nuclease-free water, Sigma-Aldrich), 20 ng/µl *Bst* LF DNA polymerase, 7.5 ng/µl HIV-1 reverse transcriptase, 0.025 ng/µl BMTU UDG and nuclease-free water to a volume of 16 µl, reserving 4 µl for sample input volume to a total of 20 µl final reaction volume. Inactivated crude samples or extracted RNA are added and the reaction is mixed by pipetting up and down several times. After that, reaction vessels were capped or sealed and incubated at 63°C for 35 minutes. For real time fluorescence detection, reactions were prepared in optically clear PCR plates and optically clear plate seals were used. Reactions were incubated in a real-time thermocycler at a block temperature of 63°C, lid temperature exceeding that of the block by at least 5°C and with real-time fluorescence readings taken every 60 s in the SYBR/FAM filter to capture Syto9 fluorescence. Colorimetric results of the bead-LAMP assays were evaluated immediately after incubation, and finished reactions were discarded to avoid cross-contamination risk.

Bead-LAMP reaction mix was prepared as above with double the amount of *Bst* LF DNA polymerase (40 ng/µl final concentration) omitting volume left for sample and replacing it with nuclease-free water to a total volume of 20 µl reaction mix per reaction. Bead extraction was performed in PCR strips or 96-well PCR plates in the following way. 100 µl of inactivated sample or extracted RNA was mixed with 60 µl of RNAClean XP beads diluted 1:5 in bead dilution buffer (2.5 M NaCl, 10 mM Tris-HCl pH 8.0, 20% (w/v) PEG 8000, 0.05% Tween 20) and pipetted up and down several times to mix. Bead-sample mixture was incubated at room temperature for 5 minutes to allow for binding of nucleic acids to the magnetic beads. Beads were then separated from the solution by placing them on a magnet for 5 minutes at room temperature. After that, the solution was removed and beads were washed twice by adding 150 µl of 85% ethanol over the beads and removing it after no longer than 30 seconds, keeping the samples on the magnet the whole time. Remaining ethanol was removed, and beads were left to air dry for a maximum of 3 minutes to avoid overdrying. 20 µl of bead-LAMP reaction mix was directly added to the beads and beads were resuspended in the mix by pipetting up and down or sealing the plate and gently vortexing. Sealed or capped reaction vessels were incubated at 63°C for 35 minutes. For real time fluorescence detection, reactions were prepared in optically clear PCR plates and optically clear plate seals were used. Reactions were incubated in a real-time thermocycler at a block temperature of 63°C, lid temperature exceeding that of the block by at least 5°C and with real-time fluorescence readings being taken every minute in the SYBR/FAM filter to capture Syto9 fluorescence. Colorimetric results of the bead-LAMP assays were evaluated immediately after incubation, and reactions were discarded.

#### Quick inactivation of sample material to bypass RNA extraction

Naso-oropharyngeal swabs collected in viral transport medium (VTM) and gargle samples collected in 5 ml 0.9% NaCl solution (saline) with roughly the same viral load were selected for RNA stability over time experiment. The samples were heat-inactivated by incubation at 95°C for 5 minutes after the addition of an inactivation reagent. The tested inactivation reagents were: QuickExtract™ DNA Extraction Solution (used as 2X solution); no reagent; 25 mM Tris(2-carboxyethyl)phosphine hydrochloride solution, pH 7.0 (TCEP, used as 10X solution); 25 mM TCEP, 10 mM EDTA (TCEP/EDTA, used as 10X solution); 25 mM TCEP, 10 mM EDTA, 4 M betaine (T/E/B, used as 10X solution); Complete inactivation solution: 25 mM TCEP, 10 mM EDTA, 4 M betaine, 2 mg/ml proteinase K (used as 10X solution). Samples were put on ice and immediately measured with direct-input qPCR in two technical duplicates as per Kellner et al., 2021, 2022 using the CDC N1 assay. Samples were split into three groups and stored at room temperature, 4°C and - 20°C. Samples were tested after 4- and 16-hours post-inactivation with direct-input RT-qPCR as described above. Delta Ct values were calculated from the geometric mean of the technical duplicates and plotted for each inactivation reagent into separate plots consisting of temperature and biological sample condition.

To demonstrate wide sample compatibility with the complete home-made and QuickExtract™ inactivation reagents, 10 SARS-CoV-2 positive nasopharyngeal swabs collected in VTM and 10 SARS-CoV-2 positive gargle samples collected in saline were inactivated with QuickExtract™ (used as 2X solution) and complete home-made (used as 10X solution) by incubation at 95°C for 5 minutes. The samples were placed on ice and immediately tested via direct-input RT-qPCR (as described in Kellner et al., 2022) to quantitatively assess the viral load, and the Ct values were compared to those obtained from these same samples with the gold-standard diagnostic RNA extracted RT-qPCR method performed at the Austrian Agency for Health and Food Safety (AGES).

#### Lyophilization of open-source RT-LAMP enzymes

Open-source RT-LAMP enzymes (*Bst* LF, HIV-1 RT and BMTU UDG) were expressed and purified as stated in this study, with a final storage buffer excluding glycerol. Protein concentration was determined using a DeNovix DS-11 FX+ Spectrophotometer/Fluorometer using calculated extinction coefficients and molecular weight values and confirmed via SDS-PAGE gel electrophoresis.

To prepare freeze-dried RT-LAMP enzyme mix, an enzyme lyophilization mixture consisting of 1x Isothermal Amplification Buffer (20 mM Tris-HCl, 10 mM (NH_4_)_2_SO_4_, 50 mM KCl, 2 mM MgSO_4_, 0.1% Tween® 20), 10% trehalose (w/v), 0.8 µg/µl *Bst* LF and 0.3 µg/µl HIV-1 RT was prepared on ice, aliquoted into 1.5 ml Eppendorf tubes with pierced caps and flash frozen in liquid nitrogen for 5 minutes. Tubes were immediately placed in a Christ Alpha 2 – 4 LDplus freeze-drier set to main drying program with chamber temperature at -78°C and pressure at 0.091 mbar. Tubes with lyophilized enzyme mixtures were collected after 16h of freeze-drying, capped with unpierced, normal lids and stored in sealable plastic bags with silica gel desiccant packets to protect the contents of the tubes from atmospheric moisture during storage.

To prepare freeze-dried LAMP reagents, a lyophilization mix was prepared consisting of 0.8X Isothermal Amplification Buffer (prepared as above, excluding MgSO_4_), 7 mM dATP, dUTP, dTTP and dCTP each, 3.5 mM dUTP, 1 µM F3/B3 primers, 2 µM LF/LB primers and 8 µM FIP/BIP primers, 100 ng/µl *Bst* LF DNA polymerase, 37.5 ng/µl HIV-1 RT, 0.125 ng/µl BMTU 3346 UDG and 10% trehalose, adding nuclease-free water to a final volume of 4 µl lyophilization mix per 20 µl reaction. This mixture was lyophilized and stored in the same way as detailed above for RT-LAMP enzymes.

Freeze-dried enzyme mixes were reconstituted by adding an amount of nuclease-free water to reach the volume of the lyophilized mix before freeze-drying. The white pellet dissolved immediately forming a ready-to-use 20X RT-LAMP enzyme mix. If storage of reconstituted enzyme mixes at -20°C is desired, reconstitution is performed with 10% glycerol in nuclease-free water instead. Reactions with reconstituted enzymes were assembled as normal RT-LAMP reactions with reconstituted enzymes being added at 1X final concentration instead of normal enzyme addition. Stability of freeze-dried LAMP enzyme mixes was tested by lyophilizing several aliquots of enzyme mix and storing them at -20°C, 4°C and room temperature to assess temperature-dependent stability. RT-LAMP reactions were assembled as described above after 0, 10 and 30 days after lyophilizing of enzyme mixes. Reactions were tested for sensitivity on a dilution series of SARS-CoV-2 positive samples with reactions assembled with non-lyophilized enzymes as a control group. Reactions were performed in 4 replicates.

Freeze-dried RT-LAMP reagent mixes were reconstituted with the addition of a reconstitution buffer that is composed of nuclease-free water, 1X Isothermal Amplification Buffer, 8 mM MgSO4, 0.4 M betaine, 120 µM HNB dye and 2 µM SYTO-9 fluorescent dye. The white pellet was readily dissolved, and the resultant mixture formed a complete RT-LAMP reaction mix that was distributed among PCR strips or PCR plates, dispensing 16 µl per well. After this, 4 µl inactivated sample was added, mixed by pipetting up and down and reactions were incubated at 63°C for 35 minutes. Stability of freeze-dried RT-LAMP reagent mixes was tested by lyophilizing several aliquots of RT-LAMP reaction mix and storing them at -20°C, 4°C, room temperature and 37°C to assess temperature-dependent stability. RT-LAMP reactions were assembled as described above after 0, 10 and 30 days after lyophilizing of enzyme mixes. Reactions were tested for sensitivity on a dilution series of SARS-CoV-2 positive samples with reactions assembled with non-lyophilized enzymes as a control group. Reactions were performed in duplicates.

#### Clinical sample collection and ethics

Patient samples in Vienna (oro/nasopharyngeal swabs and gargle) were obtained as part of a clinical performance study approved by the local Ethics Committee of the City of Vienna (#EK 20-292-1120). Oro/Nasopharyngeal swabs were collected in 3 ml Viral Transport Medium (VTM) or 0.9% NaCl solution (saline). Gargle samples were collected from patients by letting individuals gargle for 1 minute with 5 ml of Hank’s Balanced Salt Solution (HBSS) or 0.9% saline solution. Informed consent was obtained from all patients.

In Ghana, sputum samples were obtained as part of a larger COVID-19 surveillance study from individuals who tested for COVID-19 at the West African Centre for Cell Biology of Infectious Pathogens (WACCBIP), University of Ghana, during the pandemic (2021). An ethical approval was obtained from the Ghana Health Service Ethics Review Committee (GHS-ERC 011/03/20). A written informed consent was obtained from each participant before enrollment. The sputum samples were obtained in 50 mL Falcon tubes and stored at -80℃ until ready for use.

#### Lyophilized open-source RT-LAMP validation in Vienna, Austria

A total of 192 de-identified respiratory tract specimens were obtained from the Austrian Agency for Health and Food Safety and heat-inactivated using the home-made complete inactivation solution. Samples were stored at -80°C before use.

Direct RT-qPCR was performed using the Luna Universal One-Step RT-qPCR kit (NEB), 1.5 µl of reference primer/probe set CDC N1 (IDT), 0.4 µl of Antarctic Thermolabile UDG (NEB) and 2 µl of inactivated sample per 20 µl reaction. PCR program included reverse transcription at 55°C for 10 minutes, initial denaturation at 95°C for 1 minute followed by 45 cycles of 95°C for 10 s and 55°C for 30 s in a BioRad CFX qPCR cycler. For inferring viral load from direct RT-qPCR Ct values, SARS-CoV-2 RNA Synthetic Control 2 (1 000 000 copies/µl, Twist Bioscience) was diluted in heat-inactivated 0.9% NaCl. 10 000, 1 000 and 100 copies/µl dilutions were performed and tested by the protocol above in duplicated. The mean Ct values were linearized and used to construct an equation via linear regression **(Supplementary Figure 4A**), which was used to relate Ct values into copies/µl. Lyophilized open-source RT-LAMP mix was prepared as stated above and reconstituted for use in the experiment immediately after lyophilization. Real-time fluorescence detection as well as colorimetric detection after 35-minute incubation was performed, the colorimetric results were scored by the researcher immediately after incubation using the colorimetry.net web app.

#### Lyophilized open-source RT-LAMP validation in Accra, Ghana

A total of 200 cryopreserved sputum samples (80 SARS-CoV-2 positives and 120 SARS-CoV-2 negatives) were selected from a previously analyzed sample pool. RNA was purified from 180 µl of sample using the QIAamp Viral RNA Mini Kit (QIAGEN) following instructions from the manufacturer. The purified RNA was stored at -80℃ until use. RT-qPCR was performed on the QuantStudio 5 system (ThermoFisher) using the Luna Universal One-Step RT-qPCR Kit (NEB). All reactions were performed in a total volume of 15 µl consisting of 1X Luna Universal One-Step Reaction Mix, 0.75 µL of Luna WarmStart RT Enzyme Mix, 0.1 µM of each of the forward and reverse primers and 2 µL of template RNA. Previously reported forward (5’- GCGTTCTTCGGAATGTCG - 3’) and reverse (5’- TTGGATCTTTGTCATCCAATTTG - 3’) primer set targeting the N-gene was used (Chan et al., 2020). The cycling conditions consisted of 15 minutes at 55℃ and an initial denaturation of 2 minutes at 95℃ followed by 40 cycles of 15 seconds at 95℃ and 60 seconds at 60℃. The specificity of the resulting amplicons was determined by using the melting curve. RT-LAMP was performed using lyophilized RT-LAMP reagent mix shipped from Vienna, Austria. The mix was reconstituted on site and extracted RNA as well as heat-inactivated samples were tested by incubating RT-LAMP reactions at 63°C for 35 minutes in a real-time thermocycler, with fluorescence readings taken in the FAM/SYBR spectrum every 60 s. The colorimetric results were scored by the researcher immediately after incubation using the colorimetry.net web app.

## Data Availability

The original contributions presented in the study are included in the article/Supplementary Material. Further inquiries can be directed to the corresponding authors.

https://www.rtlamp.org/

## ETHICS STATEMENT

The studies involving human participants in Vienna were reviewed and approved by the local Ethic Committee of Vienna, Austria (#EK 20-208-0920). The patients/participants provided their written informed consent to participate in this study. For sample collection in Ghana, an ethical approval was obtained from the Ghana Health Service Ethics Review Committee (GHS-ERC 011/03/20). Additional written informed consent was obtained from all participants before enrolment.

## AUTHOR CONTRIBUTIONS

M.M. and M.J.K. designed, performed and analyzed all experiments conducted in Vienna; F.A. performed and analyzed experiments conducted in Ghana. I.G. performed protein expression and purification. D.H and R.H participated in experiments involving bead-LAMP. B.B. conducted cloning. L.M.-A. provided an HIV-RT expression plasmid and assisted in designing expression and purification protocols. T.O.A. and L.L.P.-G. were involved in initiating the collaborative project with Ghana, and contributed to project discussions and project funding together with J.B. and A.P. J.M.P. supported the project with providing lab space for the team and continuous funding for M.K. The VCDI enabled and supported COVID-19 testing initiatives at the Vienna BioCenter. G.A.A. supervised experiments conducted in Ghana. J.B. and A.P. conceived, coordinated and supervised the project. M.M. and M.J.K., with the help of F.A., wrote the original draft of the paper; M.M., M.J.K., J.B. and A.P. revised the paper with input from all authors.

## ACKNOWLEDGMENTS

This work would not have been possible without the enthusiastic support of the IMBA, IMP and WACCBIP research institutes, as well as the many volunteers and partners of the VCDI, who came together to help and collaborate under the exceptional circumstances of the COVID-19 pandemic. We thank the Pauli, Brennecke and Penninger groups for sharing lab space and reagents. We thank Nathan Tanner (NEB) for valuable discussions, sharing important information on LAMP technology and feedback on the manuscript, Stuart Le Grice and Jennifer Miller (NIH/NCI) for helpful advice and reagents for expression of HIV-RTs, and Jennifer C. Molloy (University of Cambridge) for useful discussion. We thank TReND in Africa for initiating the collaboration between our labs in Vienna and Ghana, and Jochen Wittbrodt, Ute Volbehr, Kevin Urbansky, and Eva Hasel for administering the VW funding. We also thank Kim and Anna Nasmyth for their generous support of the project via the “Mila Charitable Organisation”. We are grateful to the Covid Testing Scaleup Slack channel for openly sharing and exchanging information.

## VCDI AUTHOR LIST

Stefan Ameres, Institute of Molecular Biotechnology of the Austrian Academy of Sciences (IMBA), Vienna Biocenter (VBC), Vienna, Austria; Benedikt Bauer, Research Institute of Molecular Pathology (IMP), Vienna Biocenter (VBC), Vienna, Austria; Nikolaus Beer, Research Institute of Molecular Pathology (IMP), Vienna Biocenter (VBC), Vienna, Austria, Institute of Molecular Biotechnology of the Austrian Academy of Sciences (IMBA), Vienna Biocenter (VBC), Vienna, Austria, and Gregor Mendel Institute (GMI), Austrian Academy of Sciences, Vienna Biocenter (VBC), Vienna, Austria; Katharina Bergauer, Research Institute of Molecular Pathology (IMP), Vienna Biocenter (VBC), Vienna, Austria; Wolfgang Binder, Max Perutz Labs, Medical University of Vienna, Vienna Biocenter (VBC), Vienna, Austria; Claudia Blaukopf, Institute of Molecular Biotechnology of the Austrian Academy of Sciences (IMBA), Vienna Biocenter (VBC), Vienna, Austria; Boril Bochev, Research Institute of Molecular Pathology (IMP), Vienna Biocenter (VBC), Vienna, Austria, Institute of Molecular Biotechnology of the Austrian Academy of Sciences (IMBA), Vienna Biocenter (VBC), Vienna, Austria, and Gregor Mendel Institute (GMI), Austrian Academy of Sciences, Vienna Biocenter (VBC), Vienna, Austria; Julius Brennecke, Institute of Molecular Biotechnology of the Austrian Academy of Sciences (IMBA), Vienna Biocenter (VBC), Vienna, Austria; Selina Brinnich, Vienna Biocenter Core Facilities GmbH (VBCF), Vienna, Austria; Aleksandra Bundalo, Research Institute of Molecular Pathology (IMP), Vienna Biocenter (VBC), Vienna, Austria; Meinrad Busslinger, Research Institute of Molecular Pathology (IMP), Vienna Biocenter (VBC), Vienna, Austria; Aleksandr Bykov, Research Institute of Molecular Pathology (IMP), Vienna Biocenter (VBC), Vienna, Austria; Tim Clausen, Research Institute of Molecular Pathology (IMP), Vienna Biocenter (VBC), Vienna, Austria, and Medical University of Vienna, Vienna Biocenter (VBC), Vienna, Austria; Luisa Cochella, Research Institute of Molecular Pathology (IMP), Vienna Biocenter (VBC), Vienna, Austria; Geert de Vries, Institute of Molecular Biotechnology of the Austrian Academy of Sciences (IMBA), Vienna Biocenter (VBC), Vienna, Austria; Marcus Dekens, Research Institute of Molecular Pathology (IMP), Vienna Biocenter (VBC), Vienna, Austria; David Drechsel, Research Institute of Molecular Pathology (IMP), Vienna Biocenter (VBC), Vienna, Austria; Zuzana Dzupinkova, Research Institute of Molecular Pathology (IMP), Vienna Biocenter (VBC), Vienna, Austria, Institute of Molecular Biotechnology of the Austrian Academy of Sciences (IMBA), Vienna Biocenter (VBC), Vienna, Austria, and Gregor Mendel Institute (GMI), Austrian Academy of Sciences, Vienna Biocenter (VBC), Vienna, Austria; Michaela Eckmann-Mader, Vienna Biocenter Core Facilities GmbH (VBCF), Vienna, Austria; Ulrich Elling, Institute of Molecular Biotechnology of the Austrian Academy of Sciences (IMBA), Vienna Biocenter (VBC), Vienna, Austria; Michaela Fellner, Research Institute of Molecular Pathology (IMP), Vienna Biocenter (VBC), Vienna, Austria; Thomas Fellner, Vienna Biocenter Core Facilities GmbH (VBCF), Vienna, Austria; Laura Fin, Research Institute of Molecular Pathology (IMP), Vienna Biocenter (VBC), Vienna, Austria; Bianca Valeria Gapp, Institute of Molecular Biotechnology of the Austrian Academy of Sciences (IMBA), Vienna Biocenter (VBC), Vienna, Austria; Gerlinde Grabmann, Vienna Biocenter Core Facilities GmbH (VBCF), Vienna, Austria; Irina Grishkovskaya, Research Institute of Molecular Pathology (IMP), Vienna Biocenter (VBC), Vienna, Austria; Astrid Hagelkruys, Institute of Molecular Biotechnology of the Austrian Academy of Sciences (IMBA), Vienna Biocenter (VBC), Vienna, Austria; Bence Hajdusits, Research Institute of Molecular Pathology (IMP), Vienna Biocenter (VBC), Vienna, Austria; David Haselbach, Research Institute of Molecular Pathology (IMP), Vienna Biocenter (VBC), Vienna, Austria; Robert Heinen, Research Institute of Molecular Pathology (IMP), Vienna Biocenter (VBC), Vienna, Austria, Institute of Molecular Biotechnology of the Austrian Academy of Sciences (IMBA), Vienna Biocenter (VBC), Vienna, Austria, and Gregor Mendel Institute (GMI), Austrian Academy of Sciences, Vienna Biocenter (VBC), Vienna, Austria; Louisa Hill, Research Institute of Molecular Pathology (IMP), Vienna Biocenter (VBC), Vienna, Austria; David Hoffmann, Institute of Molecular Biotechnology of the Austrian Academy of Sciences (IMBA), Vienna Biocenter (VBC), Vienna, Austria; Stefanie Horer, Research Institute of Molecular Pathology (IMP), Vienna Biocenter (VBC), Vienna, Austria; Harald Isemann, Research Institute of Molecular Pathology (IMP), Vienna Biocenter (VBC), Vienna, Austria; Robert Kalis, Research Institute of Molecular Pathology (IMP), Vienna Biocenter (VBC), Vienna, Austria; Max Kellner, Research Institute of Molecular Pathology (IMP), Vienna Biocenter (VBC), Vienna, Austria, and Institute of Molecular Biotechnology of the Austrian Academy of Sciences (IMBA), Vienna Biocenter (VBC), Vienna, Austria; Juliane Kley, Research Institute of Molecular Pathology (IMP), Vienna Biocenter (VBC), Vienna, Austria; Thomas Köcher, Vienna Biocenter Core Facilities GmbH (VBCF), Vienna, Austria; Alwin Köhler, Max Perutz Labs, Medical University of Vienna, Vienna Biocenter (VBC), Vienna, Austria; Darja Kordic, Research Institute of Molecular Pathology (IMP), Vienna Biocenter (VBC), Vienna, Austria; Christian Krauditsch, Institute of Molecular Biotechnology of the Austrian Academy of Sciences (IMBA), Vienna Biocenter (VBC), Vienna, Austria; Sabina Kula, Research Institute of Molecular Pathology (IMP), Vienna Biocenter (VBC), Vienna, Austria, Institute of Molecular Biotechnology of the Austrian Academy of Sciences (IMBA), Vienna Biocenter (VBC), Vienna, Austria, and Gregor Mendel Institute (GMI), Austrian Academy of Sciences, Vienna Biocenter (VBC), Vienna, Austria; Richard Latham, Research Institute of Molecular Pathology (IMP), Vienna Biocenter (VBC), Vienna, Austria; Marie-Christin Leitner, Institute of Molecular Biotechnology of the Austrian Academy of Sciences (IMBA), Vienna Biocenter (VBC), Vienna, Austria; Thomas Leonard, Max Perutz Labs, Medical University of Vienna, Vienna Biocenter (VBC), Vienna, Austria; Dominik Lindenhofer, Institute of Molecular Biotechnology of the Austrian Academy of Sciences (IMBA), Vienna Biocenter (VBC), Vienna, Austria; Raphael Arthur Manzenreither, Institute of Molecular Biotechnology of the Austrian Academy of Sciences (IMBA), Vienna Biocenter (VBC), Vienna, Austria; Karl Mechtler, Research Institute of Molecular Pathology (IMP), Vienna Biocenter (VBC), Vienna, Austria; Anton Meinhart, Research Institute of Molecular Pathology (IMP), Vienna Biocenter (VBC), Vienna, Austria; Stefan Mereiter, Institute of Molecular Biotechnology of the Austrian Academy of Sciences (IMBA), Vienna Biocenter (VBC), Vienna, Austria; Thomas Micheler, Vienna Biocenter Core Facilities GmbH (VBCF), Vienna, Austria; Paul Moeseneder, Institute of Molecular Biotechnology of the Austrian Academy of Sciences (IMBA), Vienna Biocenter (VBC), Vienna, Austria; Tobias Neumann, Research Institute of Molecular Pathology (IMP), Vienna Biocenter (VBC), Vienna, Austria; Simon Nimpf, Research Institute of Molecular Pathology (IMP), Vienna Biocenter (VBC), Vienna, Austria; Magnus Nordborg,Gregor Mendel Institute (GMI), Austrian Academy of Sciences, Vienna Biocenter (VBC), Vienna, Austria; Egon Ogris, Max Perutz Labs, Medical University of Vienna, Vienna Biocenter (VBC), Vienna, Austria; Michaela Pagani, Research Institute of Molecular Pathology (IMP), Vienna Biocenter (VBC), Vienna, Austria; Andrea Pauli, Research Institute of Molecular Pathology (IMP), Vienna Biocenter (VBC), Vienna, Austria; JanMichael Peters, Research Institute of Molecular Pathology (IMP), Vienna Biocenter (VBC), Vienna, Austria, and Medical University of Vienna, Vienna Biocenter (VBC), Vienna, Austria; Petra Pjevac, Centre for Microbiology and Environmental Systems Science, University of Vienna, Vienna, Austria, and Joint Microbiome Facility of the University of Vienna and Medical University of Vienna, Vienna, Austria; Clemens Plaschka, Research Institute of Molecular Pathology (IMP), Vienna Biocenter (VBC), Vienna, Austria; Martina Rath, Research Institute of Molecular Pathology (IMP), Vienna Biocenter (VBC), Vienna, Austria; Daniel Reumann, Institute of Molecular Biotechnology of the Austrian Academy of Sciences (IMBA), Vienna Biocenter (VBC), Vienna, Austria; Sarah Rieser, Research Institute of Molecular Pathology (IMP), Vienna Biocenter (VBC), Vienna, Austria; Marianne Rocha-Hasler, Centre for Microbiology and Environmental Systems Science, University of Vienna, Vienna, Austria; Alan Rodriguez, Research Institute of Molecular Pathology (IMP), Vienna Biocenter (VBC), Vienna, Austria, and Institute of Molecular Biotechnology of the Austrian Academy of Sciences (IMBA), Vienna Biocenter (VBC), Vienna, Austria; James Julian Ross, Institute of Molecular Biotechnology of the Austrian Academy of Sciences (IMBA), Vienna Biocenter (VBC), Vienna, Austria; Harald Scheuch, Research Institute of Molecular Pathology (IMP), Vienna Biocenter (VBC), Vienna, Austria, Institute of Molecular Biotechnology of the Austrian Academy of Sciences (IMBA), Vienna Biocenter (VBC), Vienna, Austria, and Gregor Mendel Institute (GMI), Austrian Academy of Sciences, Vienna Biocenter (VBC), Vienna, Austria; Karina Schindler, Research Institute of Molecular Pathology (IMP), Vienna Biocenter (VBC), Vienna, Austria; Clara Schmidt, Institute of Molecular Biotechnology of the Austrian Academy of Sciences (IMBA), Vienna Biocenter (VBC), Vienna, Austria; Hannes Schmidt, Centre for Microbiology and Environmental Systems Science, University of Vienna, Vienna, Austria; Jakob Schnabl, Institute of Molecular Biotechnology of the Austrian Academy of Sciences (IMBA), Vienna Biocenter (VBC), Vienna, Austria; Stefan Schüchner,MaxPerutzLabs, Medical University of Vienna, Vienna Biocenter (VBC), Vienna, Austria; Tanja Schwickert, Research Institute of Molecular Pathology (IMP), Vienna Biocenter (VBC), Vienna, Austria; Andreas Sommer, Vienna Biocenter Core Facilities GmbH (VBCF), Vienna, Austria; Johannes Stadlmann, Institute of Biochemistry, University of Natural Resources and Life Sciences (BOKU), Vienna, Austria; Alexander Stark, Research Institute of Molecular Pathology (IMP), Vienna Biocenter (VBC), Vienna, Austria, and Medical University of Vienna, Vienna Biocenter (VBC), Vienna, Austria; Peter Steinlein, Research Institute of Molecular Pathology (IMP), Vienna Biocenter (VBC), Vienna, Austria, Institute of Molecular Biotechnology of the Austrian Academy of Sciences (IMBA), Vienna Biocenter (VBC), Vienna, Austria, and Gregor Mendel Institute (GMI), Austrian Academy of Sciences, Vienna Biocenter (VBC), Vienna, Austria; Simon Strobl, Vienna Biocenter Core Facilities GmbH (VBCF), Vienna, Austria; Qiong Sun, Research Institute of Molecular Pathology (IMP), Vienna Biocenter (VBC), Vienna, Austria; Wen Tang, Research Institute of Molecular Pathology (IMP), Vienna Biocenter (VBC), Vienna, Austria; Linda Trübestein, Max Perutz Labs, Medical University of Vienna, Vienna Biocenter (VBC), Vienna, Austria; Christian Umkehrer, Research Institute of Molecular Pathology (IMP), Vienna Biocenter (VBC), Vienna, Austria; Sandor Urmosi-Incze, Vienna Biocenter Core Facilities GmbH (VBCF), Vienna, Austria; Kristina Uzunova, Research Institute of Molecular Pathology (IMP), Vienna Biocenter (VBC), Vienna, Austria, Institute of Molecular Biotechnology of the Austrian Academy of Sciences (IMBA), Vienna Biocenter (VBC), Vienna, Austria, and Gregor Mendel Institute (GMI), Austrian Academy of Sciences, Vienna Biocenter (VBC), Vienna, Austria; Gijs Versteeg, Department of Microbiology, Immunobiology, and Genetics, Max Perutz Labs, University of Vienna, Vienna Biocenter (VBC), Dr. Bohr-Gasse 9, 1030 Vienna, Austria; Alexander Vogt, Vienna Biocenter Core Facilities GmbH (VBCF), Vienna, Austria; Vivien Vogt, Research Institute of Molecular Pathology (IMP), Vienna Biocenter (VBC), Vienna, Austria; Michael Wagner, Centre for Microbiology and Environmental Systems Science, University of Vienna, Vienna, Austria, and Joint Microbiome Facility of the University of Vienna and Medical University of Vienna, Vienna, Austria; Martina Weissenboeck, Research Institute of Molecular Pathology (IMP), Vienna Biocenter (VBC), Vienna, Austria; Barbara Werner, Vienna Biocenter Core Facilities GmbH (VBCF), Vienna, Austria; Ramesh Yelagandula, Institute of Molecular Biotechnology of the Austrian Academy of Sciences (IMBA), Vienna Biocenter (VBC), Vienna, Austria; Johannes Zuber, Research Institute of Molecular Pathology (IMP), Vienna Biocenter (VBC), Vienna, Austria, and Medical University of Vienna, Vienna Biocenter (VBC), Vienna, Austria.

## Supplementary Figures

**Supplementary Figure 1:**
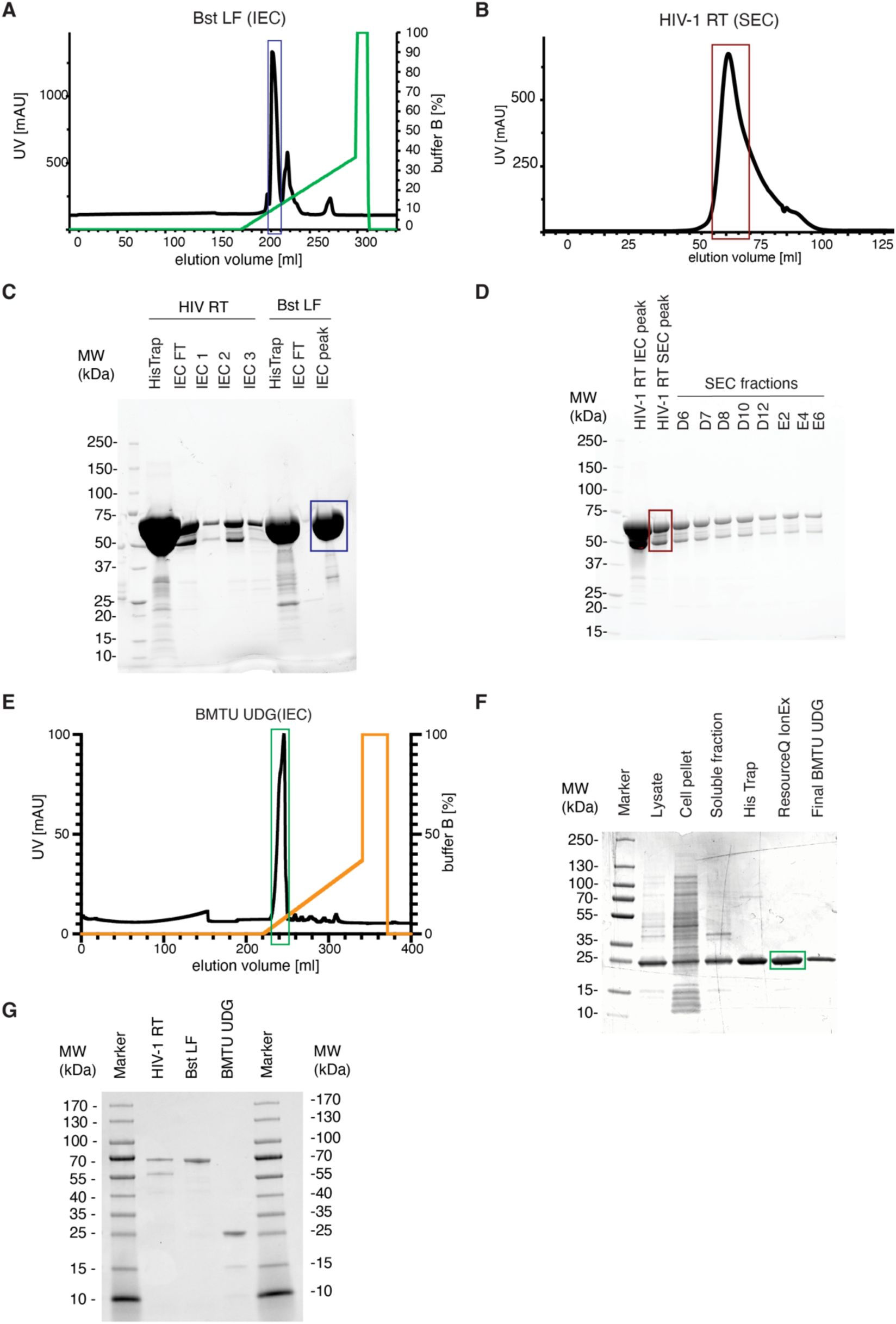
Purification of open-source enzymes (related to Figure 1) (A) Chromatograph of ion exchange purification step of *Bst* LF. Ion exchange chromatography (IEC) was performed on Resource Q 6 ml ion exchange column. Absorbance measured at 280 nm in milli-arbitrary units (mAU) is plotted against elution volume in milliliters. (B) Chromatograph of size exclusion purification step of HIV-1 RT. Size exclusion chromatography (SEC) was performed on HiLoad 16/600 Superdex 200 pg column. Absorbance measured at 280 nm in milli-arbitrary units (mAU) is plotted against elution volume in milliliters. The dark red box corresponds to the protein fractions that were pooled for use in downstream assays and is visualized in the box of the same color in SDS-PAGE gel image in Supplementary Figure 1 – D. (C) Coomassie-stained SDS-PAGE image of HIV-1 RT and *Bst* LF purification fractions. SDS-PAGE was run on the fractions of multi-step purifications of HIV-1 RT and *Bst* LF. The dark blue box corresponds to the Bst-LF protein fractions that were pooled from A) and used in downstream assays. (D) Coomassie-stained SDS-PAGE image of HIV-1 RT size exclusion chromatography purification fractions. Protein fractions collected during size exclusion chromatography purification of HIV-1 RT were loaded onto SDS-PAGE. Note the heterodimeric nature of HIV-1 RT. The dark red box corresponds to the HIV-RT protein fractions that were pooled from B) and used in downstream assays. (E) Chromatograph of ion exchange purification step of BMTU UDG. Ion exchange chromatography was performed on Resource Q 6 ml ion exchange column. Absorbance measured at 280 nm in milli-arbitrary units (mAU) is plotted against elution volume in milliliters. (F) SDS-PAGE image of BMTU UDG purification fractions. Pooled protein containing fractions from Ni-NTA and ion exchange chromatography are marked as “His Trap” and “ResQ IonEx”, respectively. Final BMTU UDG lane contains the purified, concentrated protein after dialysis. The dark green box corresponds to the BMTU UDG protein fractions that were pooled from E) and used in downstream assays. (G) SDS-PAGE image of in-house purified open-access RT-LAMP proteins. HIV-1 RT, *Bst* LF and BMTU UDG enzymes were purified as described in this study. Approximately 1.25 µg of total protein was loaded per lane.

**Supplementary Figure 2:**
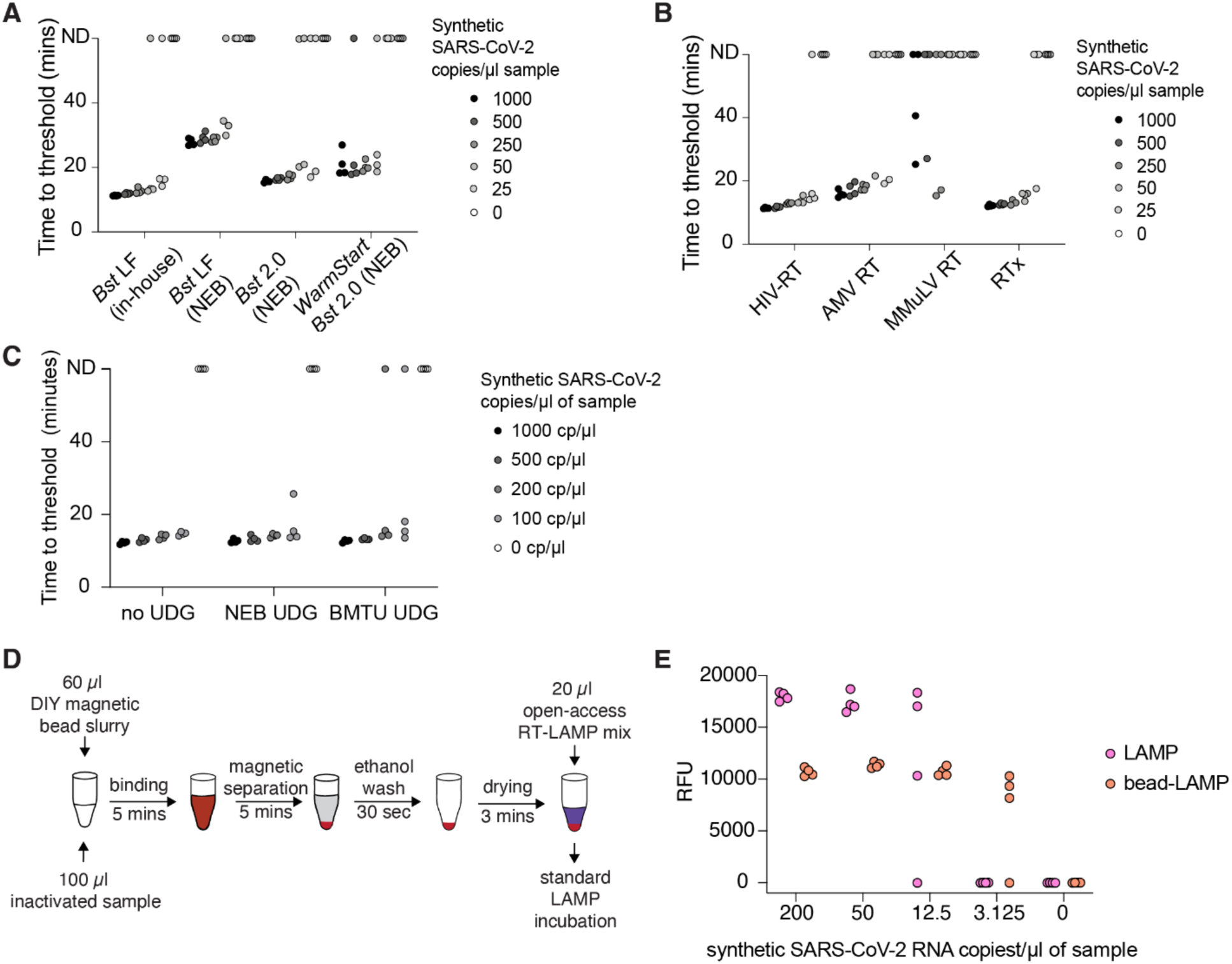
Benchmarking of open-source RT-LAMP reagents (related to Figure 1) (A) Time to threshold for DNA polymerase enzyme comparison in RT-LAMP. Time to threshold data for experiment shown in Figure 1 – A. Four replicates per condition were performed. (B) Time to threshold for reverse transcriptase enzyme comparison in RT-LAMP. Time to threshold data for experiment shown in Figure 1 – B. Four replicates per condition were performed. (C) Time to threshold for sensitivity test of RT-LAMP reactions with UDG enzymes. Reactions were prepared with *Bst* LF and HIV-1 RT. In-house BMTU UDG, Antarctic thermolabile UDG or no UDG enzymes were included. Four replicates per condition were performed. (D) Schematic representation for bead-LAMP workflow. Bead-LAMP was performed according to the protocol published previously (Kellner, Ross et al. 2022). In brief, 100 µl of sample (TCEP/Betaine/ProteinaseK-inactivated sample) was mixed with 60 µl of magnetic bead slurry and left to incubate for 5 minutes to facilitate binding of nucleic acids to magnetic beads. The beads were separated on a magnet for 5 minutes or until the solution turned completely clear. Supernatant was discarded and 200 µl of 85% ethanol was added. After 30 seconds, the ethanol was removed, and beads were left to dry out in the tube for a period of 3 minutes. RT-LAMP reaction mix was added directly on top of the dried beads and beads were resuspended in the reaction mix. The reaction vessel was capped/sealed and incubated for 35 minutes at 63°C. (E) Bead-LAMP increases sensitivity of open-source RT-LAMP reactions. Samples were prepared by diluting synthetic SARS-CoV-2 RNA in inactivated negative gargle sample in a serial dilution. The samples were tested by RT-LAMP and bead-LAMP in parallel to demonstrate the sensitivity boost provided by the simple bead-enrichment protocol.

**Supplementary Figure 3:**
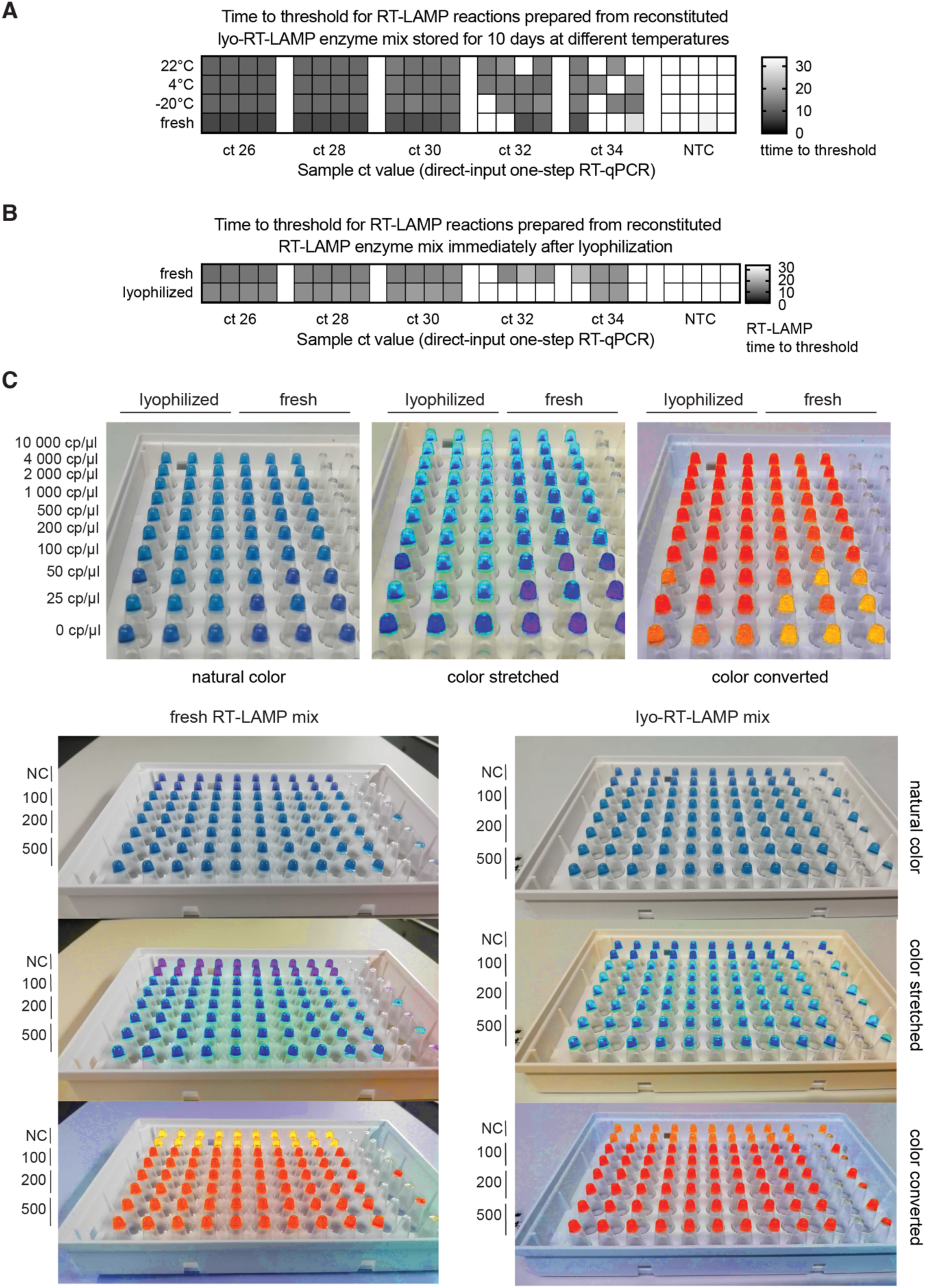
Assessment of performance of lyophilized RT-LAMP reaction mixes (related to Figure 3) (A) Performance of RT-LAMP reactions assembled with reconstituted RT-LAMP enzyme mix stored at different temperature conditions for 10 days. RT-LAMP enzyme mix was lyophilized and stored at 22°C, 4°C and -20°C. After 10 days, enzymes mixes were reconstituted in nuclease-free water and used to assemble RT-LAMP reactions, with a control of cold-stored enzymes. Reactions were tested on a four-fold, five-step dilution series of inactivated positive sample in four replicates. Direct-input RT-qPCR Ct values are displayed under the columns. (B) Performance of RT-LAMP reactions assembled with reconstituted RT-LAMP enzyme mix 0 days after lyophilization. RT-LAMP enzyme mix was lyophilized and tested immediately after lyophilization and compared to cold-stored enzymes. Reactions were tested on a four-fold, five-step dilution series of inactivated positive sample. Direct-input RT-qPCR Ct values are displayed under the columns. (C) Sensitivity comparison of lyophilized vs non-lyophilized RT-LAMP reactions on a dilution series of synthetic SARS-CoV-2 RNA (top). Colorimetric readout of reactions presented in Figure 3 – D (bottom). Smartphone images were taken after a 35-minute incubation of RT-LAMP reactions at 63°C. Color-stretched and color-converted images were generated using colorimetry.net. A notable shift in color is observed with lyophilized reagents, but reactions can still be robustly read-out based on colorimetry alone.

**Supplementary Figure 4:**
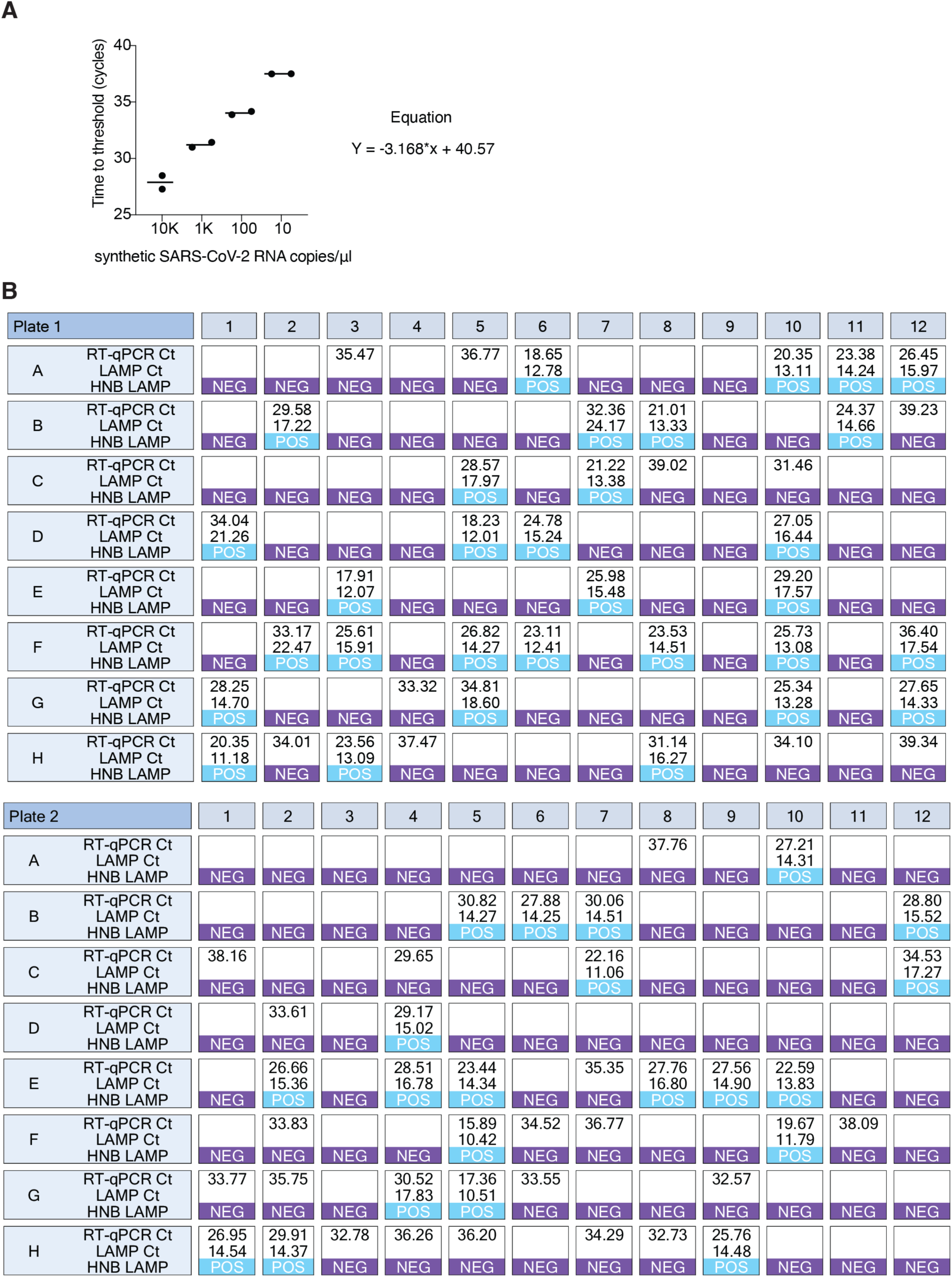
Agreement between RT-LAMP and RT-qPCR on clinical samples (related to Figure 4) (A) Dilution series of synthetic SARS-CoV-2 RNA in TCEP/Betaine/ProteinaseK-inactivated saline used for converting Ct values obtained on clinical samples into viral load in copies/µl. Synthetic SARS-CoV-2 dilutions were prepared in TCEP/Betaine/ProteinaseK-inactivated negative saline as the dilution matrix. Defined dilutions were tested in replicates and mean values of these measurements were used to generate an equation translating Ct value to copies/µl. (B) Correspondence of RT-LAMP and RT-qPCR. Shown are RT-qPCR Ct-values with corresponding RT-LAMP time-to-threshold values and HNB RT-LAMP colorimetric results for samples shown in Figure 4B.

## REFERENCES

Alekseenko, A., Barrett, D., Pareja-Sanchez, Y., Howard, R. J., Strandback, E., Ampah-Korsah, H., Rovšnik, U., Zuniga-Veliz, S., Klenov, A., Malloo, J., Ye, S., Liu, X., Reinius, B., Elsässer, S. J., Nyman, T., Sandh, G., Yin, X., & Pelechano, V. (2021). Direct detection of SARS-CoV-2 using non-commercial RT-LAMP reagents on heat-inactivated samples. Scientific Reports 2021 *11*:1, *11*(1), 1–10. 10.1038/s41598-020-80352-8

Barnes, W. M., Zhang, Z., & Kermekchiev, M. B. (2021). A Single Amino Acid Change to Taq DNA Polymerase Enables Faster PCR, Reverse Transcription and Strand-Displacement. Frontiers in Bioengineering and Biotechnology, 8, 553474. 10.3389/FBIOE.2020.553474/BIBTEX

Bhadra, S., Pothukuchy, A., Shroff, R., Cole, A. W., Byrom, M., Ellefson, J. W., Gollihar, J. D., & Ellington, A. D. (2018). Cellular reagents for diagnostics and synthetic biology. PLOS ONE, 13(8), e0201681. 10.1371/JOURNAL.PONE.0201681

Broughton, J. P., Deng, X., Yu, G., Fasching, C. L., Servellita, V., Singh, J., Miao, X., Streithorst, J. A., Granados, A., Sotomayor-Gonzalez, A., Zorn, K., Gopez, A., Hsu, E., Gu, W., Miller, S., Pan, C. Y., Guevara, H., Wadford, D. A., Chen, J. S., & Chiu, C. Y. (2020). CRISPR–Cas12-based detection of SARS-CoV-2. Nature Biotechnology 2020 *38*:7, *38*(7), 870–874. 10.1038/s41587-020-0513-4

Carter, C., Akrami, K., Hall, D., Smith, D., & Aronoff-Spencer, E. (2017). Lyophilized visually readable loop-mediated isothermal reverse transcriptase nucleic acid amplification test for detection Ebola Zaire RNA. Journal of Virological Methods, 244, 32. 10.1016/J.JVIROMET.2017.02.013

Feddema, J. J., Fernald, K. D. S., Keijser, B. J. F., Kieboom, J., & Burgwal, L. H. M. van de. (2024). Commercial Opportunity or Addressing Unmet Needs—Loop-Mediated Isothermal Amplification (LAMP) as the Future of Rapid Diagnostic Testing? Diagnostics 2024, *Vol. 14, Page* 1845, *14*(17), 1845. 10.3390/DIAGNOSTICS14171845

Fomsgaard, A. S., & Rosenstierne, M. W. (2020). An alternative workflow for molecular detection of SARS-CoV-2 - Escape from the NA extraction kit-shortage, Copenhagen, Denmark, March 2020. Eurosurveillance, *25*(14), 2000398. 10.2807/1560-7917.ES.2020.25.14.2000398/CITE/REFWORKS

Goto, M., Honda, E., Ogura, A., Nomoto, A., & Hanaki, K. I. (2009). Colorimetric Detection of Loop- Mediated Isothermal Amplification Reaction by Using Hydroxy Naphthol Blue. BioTechniques, 46(3), 167–172. 10.2144/000113072

Hsieh, K., Mage, P. L., Csordas, A. T., Eisenstein, M., & Soh, H. T. (2014). Simultaneous elimination of carryover contamination and detection of DNA with uracil-DNA-glycosylase-supplemented loop- mediated isothermal amplification (UDG-LAMP). Chemical Communications (Cambridge, England), 50(28), 3747–3749. 10.1039/C4CC00540F

Jaeger, S., Schmuck, R., & Sobek, H. (2000). Molecular cloning, sequency, and expression of the heat- labile uracil-DNA glycosylase from a marine psychrophilic bacterium, strain BMTU3346. Extremophiles : Life under Extreme Conditions, 4(2), 115–122. 10.1007/S007920050145

Jani, I. V., & Peter, T. F. (2022). Nucleic Acid Point-of-Care Testing to Improve Diagnostic Preparedness. Clinical Infectious Diseases, 75(4), 723–728. 10.1093/CID/CIAC013

Ong, J., Evans, T. J., & Tanner, N. (2012). DNA polymerases (Issue US 8993298 B1). https://lens.org/115-799-077-003-657

Joung, J., Ladha, A., Saito, M., Segel, M., Bruneau, R., Huang, M.-L. W., Kim, N.-G., Yu, X., Li, J., Walker, B. D., Greninger, A. L., Jerome, K. R., Gootenberg, J. S., Abudayyeh, O. O., & Zhang, F. (2020). Point-of-care testing for COVID-19 using SHERLOCK diagnostics. MedRxiv, 2020.05.04.20091231. 10.1101/2020.05.04.20091231

Kellner, M. J., Matl, M., Ross, J. J., Schnabl, J., Handler, D., Heinen, R., Schaeffer, J., Hufnagl, P., Indra, A., Dekens, M. P. S., Fritsche-Polanz, R., Födinger, M., Zuber, J., Allerberger, F., Pauli, A., & Brennecke, J. (2021). Head-to-head comparison of direct-input RT-PCR and RT-LAMP against RT- qPCR on extracted RNA for rapid SARS-CoV-2 diagnostics. MedRxiv, 2021.01.19.21250079. 10.1101/2021.01.19.21250079

Kellner, M. J., Ross, J. J., Schnabl, J., Dekens, M. P. S., Matl, M., Heinen, R., Grishkovskaya, I., Bauer, B., Stadlmann, J., Menéndez-Arias, L., Straw, A. D., Fritsche-Polanz, R., Traugott, M., Seitz, T., Zoufaly, A., Födinger, M., Wenisch, C., Zuber, J., Pauli, A., & Brennecke, J. (2022). A Rapid, Highly Sensitive and Open-Access SARS-CoV-2 Detection Assay for Laboratory and Home Testing. Frontiers in Molecular Biosciences, *9*, 801309. 10.3389/FMOLB.2022.801309/BIBTEX

Leonhardt, F., Gennari, A., Paludo, G. B., Schmitz, C., da Silveira, F. X., Moura, D. C. D. A., Renard, G., Volpato, G., & Volken de Souza, C. F. (2023). A systematic review about affinity tags for one-step purification and immobilization of recombinant proteins: integrated bioprocesses aiming both economic and environmental sustainability. 3 Biotech, *13*(6), 186. 10.1007/s13205-023-03616-w

Barnes Wayne, M., Kermekchiev Milko B, & Zhang Zhian. (2021). *MUTANT POLYMERASES AND USES THEREOF* (Issue US 2021/0348142 A1). https://lens.org/081-298-581-564-608

McMahon, A. (2021). Global equitable access to vaccines, medicines and diagnostics for COVID-19: The role of patents as private governance. Journal of Medical Ethics, 47(3), 142–148. 10.1136/MEDETHICS-2020-106795

Mfuh, K. O., Abanda, N. N., & Titanji, B. K. (2023). Strengthening diagnostic capacity in Africa as a key pillar of public health and pandemic preparedness. PLOS Global Public Health, 3(6), e0001998. 10.1371/JOURNAL.PGPH.0001998

Myhrvold, C., Freije, C. A., Gootenberg, J. S., Abudayyeh, O. O., Metsky, H. C., Durbin, A. F., Kellner, M. J., Tan, A. L., Paul, L. M., Parham, L. A., Garcia, K. F., Barnes, K. G., Chak, B., Mondini, A., Nogueira, M. L., Isern, S., Michael, S. F., Lorenzana, I., Yozwiak, N. L., … Sabeti, P. C. (2018). Field-deployable viral diagnostics using CRISPR-Cas13. Science, 360(6387), 444–448. 10.1126/science.aas8836

Nkengasong, J. (2020). Let Africa into the market for COVID-19 diagnostics. Nature, 580(7805), 565–565. 10.1038/D41586-020-01265-0

Notomi, T., Okayama, H., Masubuchi, H., Yonekawa, T., Watanabe, K., Amino, N., & Hase, T. (2000). Loop-mediated isothermal amplification of DNA. Nucleic Acids Research, 28(12), e63–e63. 10.1093/NAR/28.12.E63

Oberacker, P., Stepper, P., Bond, D. M., Höhn, S., Focken, J., Meyer, V., Schelle, L., Sugrue, V. J., Jeunen, G. J., Moser, T., Hore, S. R., von Meyenn, F., Hipp, K., Hore, T. A., & Jurkowski, T. P. (2019). Bio-On-Magnetic-Beads (BOMB): Open platform for high-throughput nucleic acid extraction and manipulation. PLoS Biology, 17(1). 10.1371/JOURNAL.PBIO.3000107

Ondoa, P., Kebede, Y., Loembe, M. M., Bhiman, J. N., Tessema, S. K., Sow, A., Sall, A. A., & Nkengasong, J. (2020). COVID-19 testing in Africa: lessons learnt. The Lancet Microbe, 1(3), e103–e104. 10.1016/S2666-5247(20)30068-9

Peeling, R. W., Heymann, D. L., Teo, Y.-Y., & Garcia, P. J. (2022). Diagnostics for COVID-19: moving from pandemic response to control. The Lancet, 399(10326), 757–768. 10.1016/S0140-6736(21)02346-1

Perkins, M. D., Dye, C., Balasegaram, M., Bréchot, C., Mombouli, J.-V., Røttingen, J.-A., Tanner, M., & Boehme, C. C. (2017). Diagnostic preparedness for infectious disease outbreaks. The Lancet, 390(10108), 2211–2214. 10.1016/S0140-6736(17)31224-2

Petti, C. A., Polage, C. R., Quinn, T. C., Ronald, A. R., & Sande, M. A. (2006). Laboratory Medicine in Africa: A Barrier to Effective Health Care. Clinical Infectious Diseases, 42(3), 377–382. 10.1086/499363

Rabe, B. A., & Cepko, C. (2020). SARS-CoV-2 detection using isothermal amplification and a rapid, inexpensive protocol for sample inactivation and purification. Proceedings of the National Academy of Sciences of the United States of America, 117(39), 24450–24458. 10.1073/PNAS.2011221117/SUPPL_FILE/PNAS.2011221117.SAPP.PDF

Robinson-McCarthy, L. R., Mijalis, A. J., Filsinger, G. T., de Puig, H., Donghia, N. M., Schaus, T. E., Rasmussen, R. A., Ferreira, R., Lunshof, J. E., Chao, G., Ter-Ovanesyan, D., Dodd, O., Kuru, E., Sesay, A. M., Rainbow, J., Pawlowski, A. C., Wannier, T. M., Angenent-Mari, N. M., Najjar, D., … Church, G. M. (2021). Laboratory-Generated DNA Can Cause Anomalous Pathogen Diagnostic Test Results. Microbiology Spectrum, 9(2), e00313–21. 10.1128/SPECTRUM.00313-21

Sandri, T. L., Inoue, J., Geiger, J., Griesbaum, J. M., Heinzel, C., Burnet, M., Fendel, R., Kremsner, P. G., Held, J., & Kreidenweiss, A. (2021). Complementary methods for SARS-CoV-2 diagnosis in times of material shortage. Scientific Reports 2021 *11*:1, *11*(1), 1–8. 10.1038/s41598-021-91457-z

Schermer, B., Fabretti, F., Damagnez, M., Di Cristanziano, V., Heger, E., Arjune, S., Tanner, N. A., Imhof, T., Koch, M., Ladha, A., Joung, J., Gootenberg, J. S., Abudayyeh, O. O., Burst, V., Zhang, F., Klein, F., Benzing, T., & Müller, R.-U. (2020). Rapid SARS-CoV-2 testing in primary material based on a novel multiplex RT-LAMP assay. PLoS One, 15(11), e0238612. 10.1371/journal.pone.0238612

Scott, A. T., Layne, T. R., O’Connell, K. C., Tanner, N. A., & Landers, J. P. (2020). Comparative Evaluation and Quantitative Analysis of Loop-Mediated Isothermal Amplification Indicators. Analytical Chemistry, 92(19), 13343–13353. 10.1021/ACS.ANALCHEM.0C02666/ASSET/IMAGES/LARGE/AC0C02666_0005.JPEG

Sharma, N., Jamwal, V. L., Nagial, S., Ranjan, M., Rath, D., & Gandhi, S. G. (2024). Current status of diagnostic assays for emerging zoonotic viruses: Nipah and Hendra. Expert Review of Molecular Diagnostics, 24(6), 473–485. 10.1080/14737159.2024.2368591

Studier, F. W. (2005). Protein production by auto-induction in high density shaking cultures. Protein Expr Purif, 41(1), 207–234. 10.1016/j.pep.2005.01.016

Takayama, I., Nakauchi, M., Takahashi, H., Oba, K., Semba, S., Kaida, A., Kubo, H., Saito, S., Nagata, S., Odagiri, T., & Kageyama, T. (2019). Development of real-time fluorescent reverse transcription loop-mediated isothermal amplification assay with quenching primer for influenza virus and respiratory syncytial virus. Journal of Virological Methods, 267, 53–58. 10.1016/J.JVIROMET.2019.02.010

Tanner, N. A., Zhang, Y., & Evans, T. C. (2015). Visual detection of isothermal nucleic acid amplification using pH-sensitive dyes. BioTechniques, 58(2), 59–68. 10.2144/000114253

Tomita, N., Mori, Y., Kanda, H., & Notomi, T. (2008). Loop-mediated isothermal amplification (LAMP) of gene sequences and simple visual detection of products. Nature Protocols 2008 *3*:5, *3*(5), 877– 882. 10.1038/nprot.2008.57

Ulloa, S., Bravo, C., Parra, B., Ramirez, E., Acevedo, A., Fasce, R., & Fernandez, J. (2020). A simple method for SARS-CoV-2 detection by rRT-PCR without the use of a commercial RNA extraction kit. Journal of Virological Methods, 285, 113960. 10.1016/J.JVIROMET.2020.113960

Venkatesan, P. (2023). New licences for the COVID-19 Technology Access Pool. The Lancet Microbe, 4(12), e971. 10.1016/s2666-5247(23)00329-4

WHO, A. R. (2023). REGIONAL STRATEGY ON DIAGNOSTIC AND LABORATORY SERVICES AND SYSTEMS, 2023–2032 FOR THE WHO AFRICAN REGION. AFR/RC73/7. https://www.afro.who.int/sites/default/files/2023-08/AFR-RC73-7-Regional%20strategy%20on%20diagnostic%20and%20laboratory%20services%20and%20systems%202023-2032%20for%20the%20WHO%20African%20Region.pdf

